# Exploring the association between housing insecurity and mental health among renters: A systematic review

**DOI:** 10.1101/2023.11.01.23297773

**Authors:** Mira Talmatzky, Laura Nohr, Christine Knaevelsrud, Helen Niemeyer

**Author notes:** Corresponding Author: (MT).

## Abstract

Adverse social and economic conditions negatively impact mental health and well-being. The present systematic review is the first to investigate the association between housing insecurity and mental health outcomes among renters, with a focus on housing affordability and instability. We followed the Preferred Reporting Items for Systematic reviews and Meta-Analyses (PRISMA) guidelines. A comprehensive search was conducted in December 2022 across four databases (MEDLINE, PsycINFO, Web of Science, and ASSIA). Quantitative studies from OECD (Organisation for Economic Co-operation and Development) member countries were eligible for inclusion if they investigated housing insecurity by examining at least one independent variable related to housing affordability and/or instability, and included at least one mental health-related outcome among adult renters. Due to heterogeneity of the identified studies, we performed a narrative synthesis. Twenty-two studies met the inclusion criteria, of which 14 applied a longitudinal design, five a cross-sectional design, and three a quasi-experimental design. Among the nine studies examining housing affordability, six reported significant associations between unaffordable rent and poor mental health in low-income renters. Regarding housing instability, 12 out of 14 studies reported significant associations between unstable housing and renters’ mental health issues. Measures of housing insecurity varied, with rent-to-income ratio and forced moves being most commonly employed. Mental health outcomes focused primarily on overall mental health, well-being, and depressive symptoms, while few studies explored other mental health outcomes. The findings suggest that experiencing unaffordable or unstable housing has a negative impact on renters’ overall mental health and depressive symptoms. Housing insecurity poses a significant challenge for renters in OECD countries, highlighting the need for policymakers to implement supportive housing policies and tenure protection measures in order to improve renters’ housing security and ultimately public health.

## Introduction

### Background

Social conditions and contextual factors significantly influence the risk, development, and persistence of mental disorders and psychological distress. According to the World Health Organization (WHO), social determinants, encompassing the conditions in which individuals are born, grow, work, live, and age, contribute to severe health inequalities within contemporary societies (WHO & Calouste Gulbenkian Foundation, 2014). Housing is widely recognized as a significant social determinant influencing mental health and contributing to public health challenges (Baumgartner et al., 2023; Compton & Shim, 2015; Lund et al., 2018; Marmot & Wilkinson, 2005; Shaw, 2004; Suglia et al., 2015; Swope & Hernández, 2019). Housing insecurity is a global phenomenon, which exposes individuals to various forms of challenges, including unaffordable housing, poor housing conditions, a lack of stable tenure, and homelessness (DeLuca & Rosen, 2022; Eurostat, 2022; Prindex, 2020; United Nations, 2020). Renters, and particularly low-income renters, are especially vulnerable to these housing disadvantages (DeLuca & Rosen, 2022; Hulse et al., 2019; Routhier, 2019). Indeed, insecure housing not only compromises the basic need for shelter but also poses significant risks to mental health.

Multiple systematic reviews have explored the association between housing insecurity and mental health outcomes, providing valuable insights within this field of research. Nevertheless, all of these reviews either did not differentiate by tenure status or focused specifically on tenures other than renting, such as home ownership or homelessness. For instance, research has investigated a range of housing disadvantages, such as eviction and foreclosure (A. C. Tsai, 2015; H. Vásquez-Vera et al., 2017), financial distress of homeowners (Downing, 2016), and homelessness (Fazel et al., 2008; Hodgson et al., 2013), and their link to mental health. In a systematic review of longitudinal studies, Singh et al. (2019) found that prior exposure to housing disadvantages can have effects on mental health later in life. Moreover, the impact of living environment and neighborhood conditions, such as poor housing quality and air pollution, on psychological distress has been extensively studied in multiple systematic reviews (Rautio et al., 2018; Richardson et al., 2015; Truong & Ma, 2006). Additionally, systematic reviews have explored the relationship between housing and mental health in specific populations, such as students (Franzoi et al., 2022), children (Jelleyman & Spencer, 2008; Y. Li et al., 2021), and individuals with severe mental illnesses (Kyle & Dunn, 2008). However, to the best of our knowledge, no review to date has specifically examined the issue of housing insecurity among renters and its impact on mental health.

### Linking Housing Insecurity to Renters’ Mental Health

Housing scholars have long emphasized that housing is more than a mere physical shelter. Rather, it constitutes a social and emotional environment that significantly impacts our psychological well-being (Easthope, 2014; Hiscock et al., 2001; Hulse & Saugeres, 2008; Padgett, 2007). The acute and chronic stress resulting from exposure to social disadvantages directly and indirectly impact the development and course of mental disorders (Marmot & Wilkinson, 2005). In a systematic review of social determinants influencing mental disorders, Lund et al. (2018) identified depression, anxiety, substance abuse, suicide, psychosis, and dementia as potential outcomes associated with economic disadvantages, including housing issues. Traditionally, the literature has emphasized the (health) benefits of homeownership, while considering rental tenure as inherently negative for mental well-being due to its lack of stability and ontological security (Dupuis & Thorns, 1998; Kearns et al., 2000). Recently, however, a growing body of research has explicitly examined the composition of cohorts living in rental housing as well as the specific rental conditions that impact mental health (Acolin, 2022; Baker et al., 2013; Herbers & Mulder, 2017; Hulse & Milligan, 2014). A factor driving this shift was the sudden rise in the number of renters, particularly within the private rental sector, in highlZlincome countries traditionally dominated by homeownership, such as Australia (Hulse & Yates, 2017), the United Kingdom (UK; Bone, 2014), and the United States of America (U.S.; Crook & Kemp, 2014). Worldwide, renters face various pressures, including constrained housing markets, rising rental costs, and the associated challenges of finding affordable, stable, and suitable housing (Baumgartner et al., 2023; Bone, 2014; Clair et al., 2019; Eurostat, 2022; Routhier, 2019). These challenges have been further reinforced by recent societal upheavals such as the COVID-19 pandemic, related social and economic lockdowns, and the energy crisis (Baker, Bentley, et al., 2020; Oswald et al., 2022; Waldron, 2022). Governments have implemented a range of housing policies and programs to address the issue of housing security, including social/public housing, housing subsidies, or eviction policies (DeLuca & Rosen, 2022; Dweik & Woodhall-Melnik, 2022). However, according to DeLuca and Rosen (2022), these housing programs frequently fail to reach those who truly need them, and many eligible renters do not receive the necessary support. In brief, renters have emerged as a vulnerable group that is disproportionately affected by housing insecurity and economic crises. Against this background, the present systematic review aims to explore the association between housing insecurity and mental health outcomes among renters.

### Conceptualization of Housing Insecurity Among Renters

Housing insecurity is a multidimensional concept. Although definitions vary across scholars, commonly applied dimensions include housing affordability, quality, (in-)stability, safety, and neighborhood opportunities (Clair et al., 2019; Cox et al., 2019; DeLuca & Rosen, 2022; Swope & Hernández, 2019). Hence, exposure to housing insecurity refers to conditions and situations in which individuals face challenges related to these dimensions. Housing insecurity intersects with insecurities in other domains, such as finance, employment, family, and health, often creating a complex interplay in which these insecurities interact and mutually reinforce each other (Hulse & Saugeres, 2008). With regard to renters, secure housing involves finding and maintaining homes that meet tenants’ needs, and is influenced by factors beyond legal tenure, including societal context (e.g., rent regulations), the market (e.g., affordability), public policy (e.g., rental assistance), cultural norms, and psychosocial dimensions of security (Hulse & Milligan, 2014; Hulse et al., 2011; James et al., 2022). In this systematic review, we focus on two primary dimensions of housing insecurity among renters: *Housing unaffordability* stands out as the most extensively studied issue among renters and is the strongest standalone dimension of housing insecurity (Clair et al., 2019; Hulse & Milligan, 2014; Routhier, 2019). Affordability refers to the ability of renters to meet the costs of housing, including rent and other related expenses such as utilities and deposits, without experiencing excessive financial strain (Clair et al., 2019; Cox et al., 2019; Routhier, 2019; Swope & Hernández, 2019). The operationalization of housing affordability varies across studies (Cox et al., 2019), with common approaches being the rent-to-income ratio (e.g., “housing-cost burden”), housing-induced poverty, residual income, and subjective measures of difficulty to afford the rent (Routhier, 2019). In the case of rent-to-income ratio, the threshold for when a household is considered unaffordable varies between 30 and 50 % (Cox et al., 2019), sometimes with the added criterion that the household income is in the bottom 40 % of the national distribution, referred to as the ‘30/40’ measure (Nepal et al., 2010).

*Housing instability* poses another primary barrier to secure housing for renters (Clair et al., 2019; Cox et al., 2019; Routhier, 2019; Swope & Hernández, 2019). Stability refers to the ability of renters to remain in their housing for as long as they wish (Clair et al., 2019; Cox et al., 2019; Routhier, 2019; Swope & Hernández, 2019). Common measurement approaches for housing instability among renters include eviction and frequency of moving (Routhier, 2019). However, compared to affordability, operationalizations of instability vary more broadly across different studies, and also include issues such as overcrowding, doubling up, duration of stay, and rental arrears (Cox et al., 2019; Swope & Hernández, 2019).

### Objectives

The present study aimed to explore exposure to housing insecurity, i.e. unaffordability and instability, and its impact on mental health among renters. For this purpose, we focused on adults living in OECD (Organisation for Economic Co-operation and Development, 2023) member countries to ensure greater comparability within socioeconomic contexts. All types of rental tenure (e.g., private or social renting) were included. Studies with renter samples in housing programs and subsidies directed at improving housing affordability and/or instability were also eligible for inclusion. Using a systematic review methodology and narrative synthesis, the primary objectives of this research were to comprehensively identify relevant study reports, synthesize key findings, and propose potential areas for future investigations. We further evaluated the reporting quality of the included studies, as transparent reporting is crucial for assessing the quality of studies and their replicability (Purssell & McCrae, 2020).

## Methods

The review was conducted and reported according to the Preferred Reporting Items for Systematic Reviews and Meta-Analyses (Page et al., 2021) guidelines (PRISMA checklist provided in S1 Appendix). To synthesize the evidence, a narrative synthesis was chosen over a meta-analysis, due to the heterogeneity of the included studies regarding methodology and the applied definitions of housing insecurity. The review was not pre-registered.

### Eligibility Criteria

To be eligible for inclusion, studies needed to be quantitative primary or secondary research published in English that investigated housing insecurity by examining at least one variable related to housing affordability, such as housing cost burden or receiving rental assistance, and/or housing instability, such as frequent moves, forced moves, or overcrowding. Studies investigating housing programs or policies aimed at improving affordability and/or stability, such as rental assistance or eviction prevention programs, were also considered for inclusion. Furthermore, studies needed to include at least one mental health-related outcome, such as specific mental disorders, overall mental well-being, suicidal behavior, or treatment for mental health reasons (e.g., medication for emotional conditions). Only data points that focused on the population of adults living in rental households were eligible for inclusion, with no restriction on the type of rental tenure (e.g., private or social renting). Longitudinal studies measuring tenure status at only one time point were eligible for inclusion if they considered their population as “renters”. Studies needed to be situated in OECD countries. No restrictions were made regarding publication date or status.

Data points were excluded if they analyzed other forms of housing insecurity such as housing quality, safety, neighborhood characteristics, and homelessness. Likewise, data points examining residential satisfaction and general health outcomes were excluded. Populations other than renters, such as homeowners or homeless individuals, were not eligible. While studies examining children and adolescents were also not eligible, studies that examined adult populations while including individuals who were at least 15 years old were included in the analysis. Editorials, reviews, qualitative studies, and research with solely descriptive data were excluded.

### Search Strategy

A comprehensive search of the literature was conducted in two stages. In the first stage, four electronic databases were searched: MEDLINE (PubMed), PsycINFO (EBSCO), Web of Science Core Collections, and ASSIA (ProQuest). These databases were selected because they cover a broad range of disciplines that are relevant to housing and mental health research, such as psychology, social sciences, and public health, and also feature a wide range of document types, including dissertations and conference proceedings. The search was based on three search components:

a. the target population of renters,
b. exposure to housing insecurity (housing affordability and/or instability), and
c. mental health-related outcomes.

To identify relevant keywords, synonyms were searched for and key studies in the field were consulted. Additionally, Medical Subject Heading (MeSH) terms were used for MEDLINE and thesaurus index terms were used for PsycINFO and ASSIA. To further enhance the sensitivity of the search strategy, wildcard symbols were utilized and no search filters or limits were applied. The literature search was conducted on December 9, 2022. The detailed search syntax is provided in S2 Appendix.

In the second stage, citation tracking was used to further increase sensitivity and reduce potential bias due to unpublished studies or studies not indexed in the selected databases. Backward citation tracking involved screening titles and abstracts of the reference lists from included studies and related systematic review. Forward citation tracking was conducted from February 23 to 27, 2023, screening the titles and abstracts of all studies that cited the included studies.

### Study Selection

After exporting all search results into the reference management program Citavi (Version 6.14), duplicate records were removed. The initial screening process involved assessing the title and abstract of each record for eligibility. If a record was considered eligible or potentially eligible, its full text was retrieved and screened accordingly. Study selection was conducted hierarchically using a screening form, based on the research question and predefined inclusion and exclusion criteria. Exclusion reasons for full-text reports were documented and are provided in S3 Appendix. It was ensured that studies which used the same database were distinct, as they examined separate research questions with different mental health-related outcomes and/or variables assessing housing affordability and/or instability. The screening process was conducted by the first author, without the use of automation tools. In cases of uncertainty, discussions with the co-author H.N. were held to reach a decision. Additionally, to ensure reliability, a randomized subset of 10 % of the full texts (*N* = 12) was independently screened by the co-author L.N. and the interrater reliability was calculated using Cohen’s kappa (Cohen, 1960).

### Data Extraction and Synthesis Method

The study characteristics of the included studies were extracted by the first author and recorded in an extraction sheet, which comprises (a) the study characteristics (e.g., author and year, country, study design, information about the study population), (b) variables (exposure and outcome variables, as well as measurements), (c) statistical methods and study limitations and strengths, and (d) main findings. The main results were extracted separately for each variable assessing housing affordability and/or instability and mental health-related outcome measure. In the summary table, adjusted results including statistical significance, relevant effect measures (mostly odds ratios or mean differences) and confidence intervals were reported whenever possible. Unadjusted results were reported only when adjusted results were not available. After the initial extraction, a verification process was conducted by the co-author L.N. in a randomized subset of 30 % of the included studies (*N* = 7) by checking whether the extracted information of the included studies was accurate. Discrepancies were resolved through discussions between the authors and the extracted information was clarified and corrected as needed.

In the evidence synthesis, the main findings of the included studies were grouped and presented by outcome and predictor variables. In an initial step, predictor variables were classified into the two dimensions housing affordability and instability. If studies examined housing programs and policies like housing benefits, social housing, or eviction moratorium, categorization was determined by their principal objectives. If studies employed different samples or multiple predictor variables belonging to different categories of housing insecurity, each of them was reported separately. Outcome measures were grouped into the categories “overall mental health, well-being, and psychological distress”, “psychosocial functioning”, “symptoms of mental disorders”, and “mental health treatment”.

### Assessment of Reporting Quality

The reporting quality of cohort and cross-sectional studies was assessed using the well-established STROBE (STrengthening the Reporting of OBservational studies in Epidemiology) statement for cross-sectional and cohort studies (Elm et al., 2008). Quasi-experimental studies were assessed using the TREND (Transparent Reporting of Evaluations with Nonrandomized Designs) statement (Des Jarlais et al., 2004). The assessment was carried out by the first author.

## Evidence Synthesis

### Study Selection

The study selection process is illustrated in the PRISMA flow diagram (Fig 1). The initial search yielded a total of 1,198 potentially eligible titles (after removal of duplicates). Following title and abstract screening, 74 full-text studies were assessed for eligibility (despite multiple attempts to contact the authors, two reports could not be retrieved). Of these, 19 studies met the inclusion criteria. Based on the initial results, a subsequent step involved identifying 49 additional reports through backward and forward citation tracking. This process resulted in the inclusion of three studies. A detailed overview of the reasons for exclusion is provided in S3 Appendix. The most common reason for exclusion was that studies did not specifically analyze the association of housing insecurity and mental health among renters but rather addressed housing insecurity more broadly (e.g., statistical analyses were not separated by tenant subgroups). Overall, a total of 22 studies were included in the evidence synthesis. The interrater reliability of the study selection process was κ = 1, indicating almost perfect agreement (Cohen, 1960; Landis & Koch, 1977).

**Fig 1.**
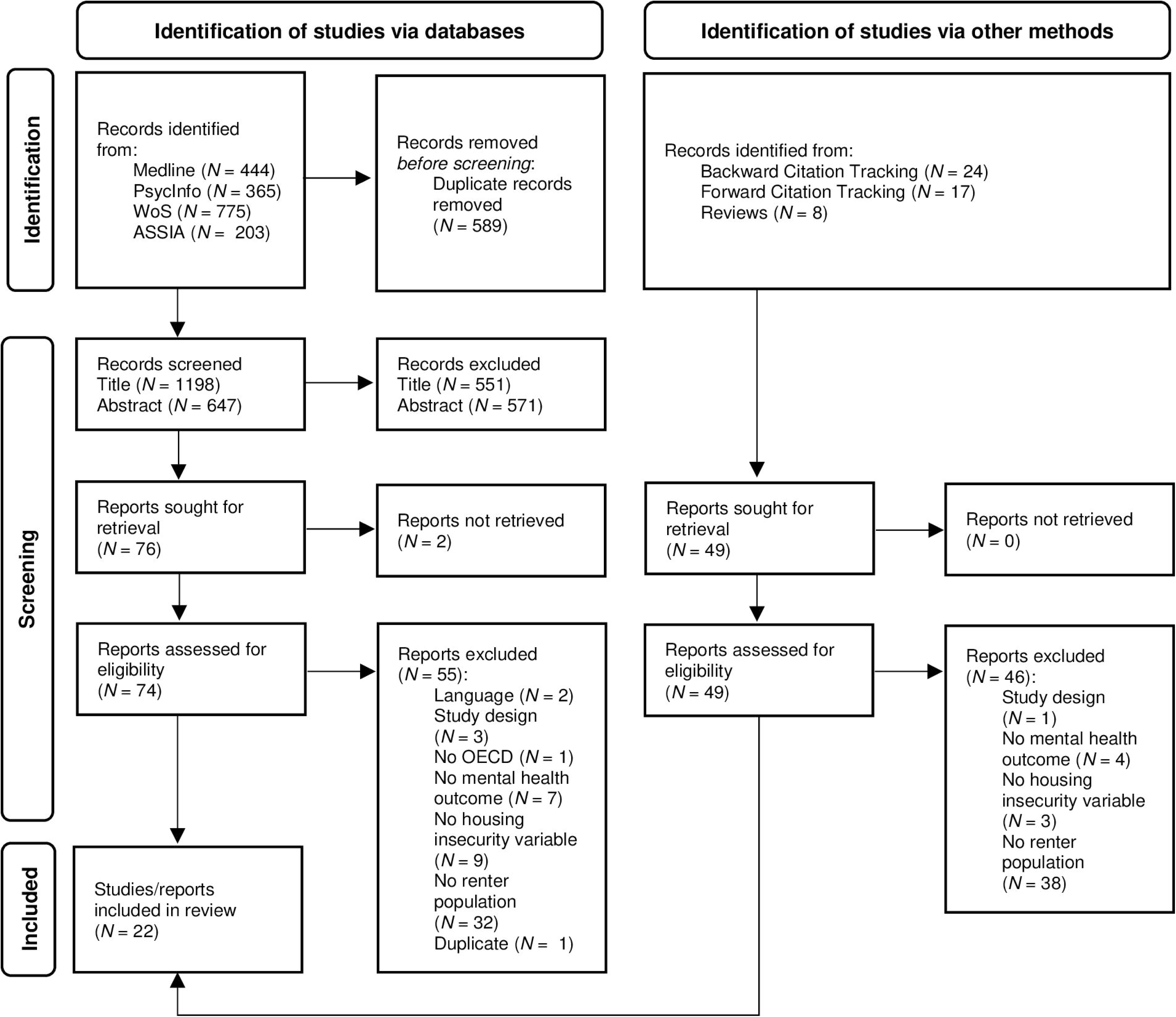
PRISMA 2020 flow diagram.

### Study Characteristics

Characteristics of the included studies are displayed in Table 1 (housing affordability) and Table 2 (housing instability). Eleven of the studies were from the U.S., six from Australia, three from the UK, and one each from the Netherlands, South Korea, and Spain. Fourteen studies were longitudinal studies, five were cross-sectional studies, and three were quasi-experimental studies without actively manipulated interventions. The majority of the studies were secondary analyses. With the exception of one study, all were published within the last decade, mostly in 2020 or later.

**Table 1.** Summary characteristics of the included studies regarding housing affordability and mental health (ordered alphabetically according to author’s name)

| Study | Country | Study Design | Data Source & Years of Collection | Main Objective of Study | Study Population (Analytical Sample) | Predictor Operationalization & Measurement<br>*Housing Insecurity* | Outcome Operationalization & Measurement<br>*Mental Health* | Main Findings |
| --- | --- | --- | --- | --- | --- | --- | --- | --- |
| **Arundel et al. (2022)** | The Netherlands | Longitudinal (Panel study) | Longitudinal Internet Studies for the Social Sciences (LISS Panel) 2008-2019 | Examining the link between housing unaffordability and mental health and how this is differentiated across age cohorts and tenure | Renters not living in parental home (complete case analysis)<br>Age 25-65 years<br>$N_{Renters} = N/A^a$ | *Housing affordability stress (HAS)*<br>'30/40' measure<sup>b</sup> | *Mental health and well-being*<br>MHI-5 from the SF-36 | Renters with HAS consistently showed lower mental health scores compared to renters without HAS, with significant differences observed at a 90 % CI, except for the years 2009 & 2012. The divergence in mental health between those with and without HAS was particularly pronounced among renters in the 35-44 age cohort. |
| **Baker et al. (2020)** | Australia | Longitudinal (Panel study) | Household, Income and Labour Dynamics in Australia (HILDA) 2002-2016 | Examining the mental health effects of (prolonged and intermittent) patterns of exposure to housing affordability problems | Private renters & public renters who took part in the survey for five consecutive waves (complete case analysis)<br>Age $\geq 15$ years<br>$N_{Private} = 1,401^c$<br>$N_{Public} = 9,694$ | *Housing affordability stress pattern*<br>Pattern of exposure to HAS over time: prolonged vs. intermittent affordability problems vs. unexposed to housing affordability | *Mental health and well-being*<br>MCS from the SF-36 | Both the intermittent and the prolonged exposure group had lower mental health scores compared to renters unexposed to HAS, with the prolonged group showing the lowest average scores.<br>For private renters, the group mean differences between prolonged vs. unexposed and between prolonged vs. intermittent HAS were not significant. For public renters, the group mean differences were all significant ($p < .01$)<sup>d</sup>. |
| **Bentley et al. (2016) [Part a]** | Australia | Longitudinal (Panel study) | HILDA 2001-2008 | Examining whether the mental health of people in different tenure types was differentially affected by unaffordable housing | Low-income private renters<br>Age 25-64 years<br>$N_{Renters} = N/A^e$<br>(3,036 observations) | *Housing cost burden*<br>Rent payments exceeded 30 % of gross household income (equivalent to '30/40' measure) | *Mental health and well-being*<br>MCS from the SF-36 | Private renters whose housing became unaffordable experienced a small but significant decline in mental health (mean change = $-0.45$; 95 % CI: $-0.81$, $-0.09$; $p = .014$). |
| **Bentley et al. (2016)**<br>[Part b] | UK | Longitudinal (Panel study) | British Household Panel Survey (BHPS)<br>2001-2008 | See [Part a] | Low-income private renters<br>Age 25-64 years<br>$N_{Renters} = N/A^1$<br>(2,373 observations) | *Housing cost burden*<br>See [Part a] | *Mental health and well-being*<br>GHQ-12 | Private renters whose housing became unaffordable experienced no significant change in mental health (mean change = 0.30; 95 % CI: -0.67, 1.26; $p = .749$). |
| **Chambers et al. (2015)**<br>[Part a] | U.S. | Cross-sectional | Affordable Housing as an Obesity Mediating Environment (AHOME)<br>2011-2012 | Investigating whether the type of housing assistance and the conditions of the home and neighborhood are associated with mental health outcomes of residents | Low-income Latin American renters living in the Bronx, eligible for rental assistance (complete case analysis)<br>Age $\geq 18$ years<br>$N = 371$ | *Receiving housing assistance*<br>Unassisted vs. public housing residents vs. Section 8 voucher | *Symptoms of depression*<br>CES-D 10<br>*Hostile affect*<br>Items of the Hostile Affect Subscale from the CMHS | Receipt of housing assistance was not significantly associated with depressive symptomology (OR = 0.857; 95 % CI: 0.48, 1.53; $p$-value N/A) or hostility (OR = 1.738; 95 % CI: 0.8, 3.76; $p$-value N/A). |
| **Elliott et al. (2021)** | U.S. | Longitudinal (Cohort study) | Fragile Families & Child Well-being Study (FFCWS)<br>1998-2017 | Analyzing the relationship between housing cost burden and mental health outcomes reported by a sample of rent-burdened primary caregiver mothers | Mothers/female caregivers from low-income rental households (complete case analysis)<br>Age N/A<br>$N = 399$ | *Housing cost burden*<br>Monthly rent as a proportion of income (< 31 % not burdened, 31-49 % burdened, > 50 % severely burdened) | *Symptoms of depression*<br>Items from the CIDI-SF | Mothers/female caregivers from low-income rental households who experienced rent burden (OR = 1.7, $p = .004$) or severe rent burden (OR = 1.5, $p = .005$) were significantly more likely to report liberal criteria for depression than those who did not experience rent burden. |
| **Mason et al. (2013)** | Australia | Longitudinal (Panel study) | HILDA<br>2001-2010 | Investigating whether a relationship exists between unaffordable housing and mental health | Low-income private renters who experienced a change in their affordability status during the study period | *Housing affordability stress (HAS)*<br>'30/40' measure | *Mental health and well-being*<br>MCS from the SF-36 | Among low-income private renters, individuals in unaffordable housing showed a mean MCS score that was 1.18 points lower than the mean score when their housing was affordable (95 % CI: 1.95, 0.41, $p = .003$). These results were consistent across different low- and mid-low-income |
| | | | | that differs between home purchasers and private renters among low-income households | Age 25-64 years<br>$N_{Renters} = N/A$<br>(4,362 observations)<sup>g</sup> | | | subgroups, but were not replicated in sensitivity analyses examining higher-income households. |
| **Prentice & Scutella (2020)** | Australia | Quasi-experimental study/longitudinal data | HILDA<sup>h</sup><br>2001-2015 | Estimating the impacts of social housing on employment, education, health, incarceration and homelessness | Renters in social housing at time t (Control group: low-income renters)<br>Age $\geq 15$ years<br>$N = N/A$ | *Living in social housing*<br>Renters in community or public housing vs. low-income renters not living in social housing (matched control group) | *Psychological distress*<br>K6<br>*Mental health and well-being*<br>MHI-5 from the SF-36 | No significant improvements in mental health arising from social housing at t+1 (6-12 months later); indeed, social housing residents had significantly worse outcomes (ATT = 1 (MHI-5), ATT = 0.6 (K6), $p < .05$). |
| **Reeves et al. (2016)** | UK | Quasi-experimental study/repeated cross-sectional data | Annual Population Survey (APS)<br>2009-2013 | Evaluating the association between housing security and mental ill health by investigating the link between a reduction in income (reduction of housing benefits) and mental health | Private renters receiving Housing Benefits (HB) (Control group: Private renters without HB)<br>Age 16-69 years<br>$N = 179,037$ | *Amount of housing benefits*<br>HB reduction in 2011 (mean reduction of approximately £1,220 per year) | *Symptoms of depression*<br>What health problems does the respondent currently have (list incl. "depression, bad nerves or anxiety") | The reduction of HB increased the risk of depressive symptoms among persons claiming the HB by approximately 1.8 percentage points (95 % CI: 1.0, 2.7; $p < .01$) above the risk in private renters not claiming the benefit. This effect remained after matching the groups. The increase in depressive symptoms remained elevated for up to 24 months after the reform. |
| **Rodgers et al. (2019)** | U.S. | Longitudinal with quasi-experimental variation (Panel study) | National Longitudinal Survey of Youths 1979 (NLSY79)<br>2000-2014 | Examining the relationship between housing affordability and risk factors for cardiovascular | Renters who were 'disease-free' prior to 2000 and experienced a change in the aggregated rent burden | *Aggregated housing cost burden*<br>Change in the county-average proportion of total household income spent on rent (before vs. after the Great | *Change in symptoms of depression*<br>CES-D (before vs. after the Great Recession 2008) | Each percentage point increase in county-level median percentage of housing cost burden was associated with increased odds of depression. The association was stronger for renters than for homeowners, although the association was no longer significant when restricting the sample to renters (OR = 1.20, 95 % CI: 0.92, 1.56; $p = .17$). |
| | | | | disease | (complete case analysis)<br>Age 35-43 years<br>$N_{Renters} = 266^i$ | Recession in 2008) | | |

Notes: *CES-D 10* Center for Epidemiologic Studies – Depression Scale; *CIDI-SF* Composite International Diagnostic Interview Short-Form; *CMHS* Cook-Medley Hostility Scale; *GHQ-12* 12-item General Health Questionnaire; *K6* Abbreviated version of Kessler Psychological Distress Scale; *MCS* Mental Health Component Summary Score; *MHI-5* Mental Health Scale; *SF-36* Short Form 36 Health Survey; References for each mental health measurement test are provided in S5 Appendix.
<sup>a</sup> $N_{Renters} = 12,306$ (before discounting cases with missing data on key variables, final sample not specified), $N_{Total} =$ about 14,000 (1,500-1,900 per year)
<sup>b</sup> '30/40' measure= spending > 30 % of gross household income on rent and gross household income is in the bottom 40 % of the national distribution (Nepal et al., 2010)
<sup>c</sup> $N_{Total} = 48,446$
<sup>d</sup> This review solely presents the descriptive findings of the study, as the regression analyses did not include stratified analyses for renters.
<sup>e</sup> $N_{Total} = 2,239$ (8,481 responses)
<sup>f</sup> $N_{Total} = 2,269$ (9,184 responses)
<sup>g</sup> $N_{Total} = 2,916$ (12,064 observations)
<sup>h</sup> The primary data source for the paper was Journey Home (JH); however, the results are not incorporated within this review, as the control group of this sample is not restricted to renters but includes all individuals not in social housing
<sup>i</sup> $N_{Total} = 3,722$ , of which $N_{Renters} = 1,117$ ; final sample size $N_{Total} = 713$ for depressive symptoms

**Table 2.**
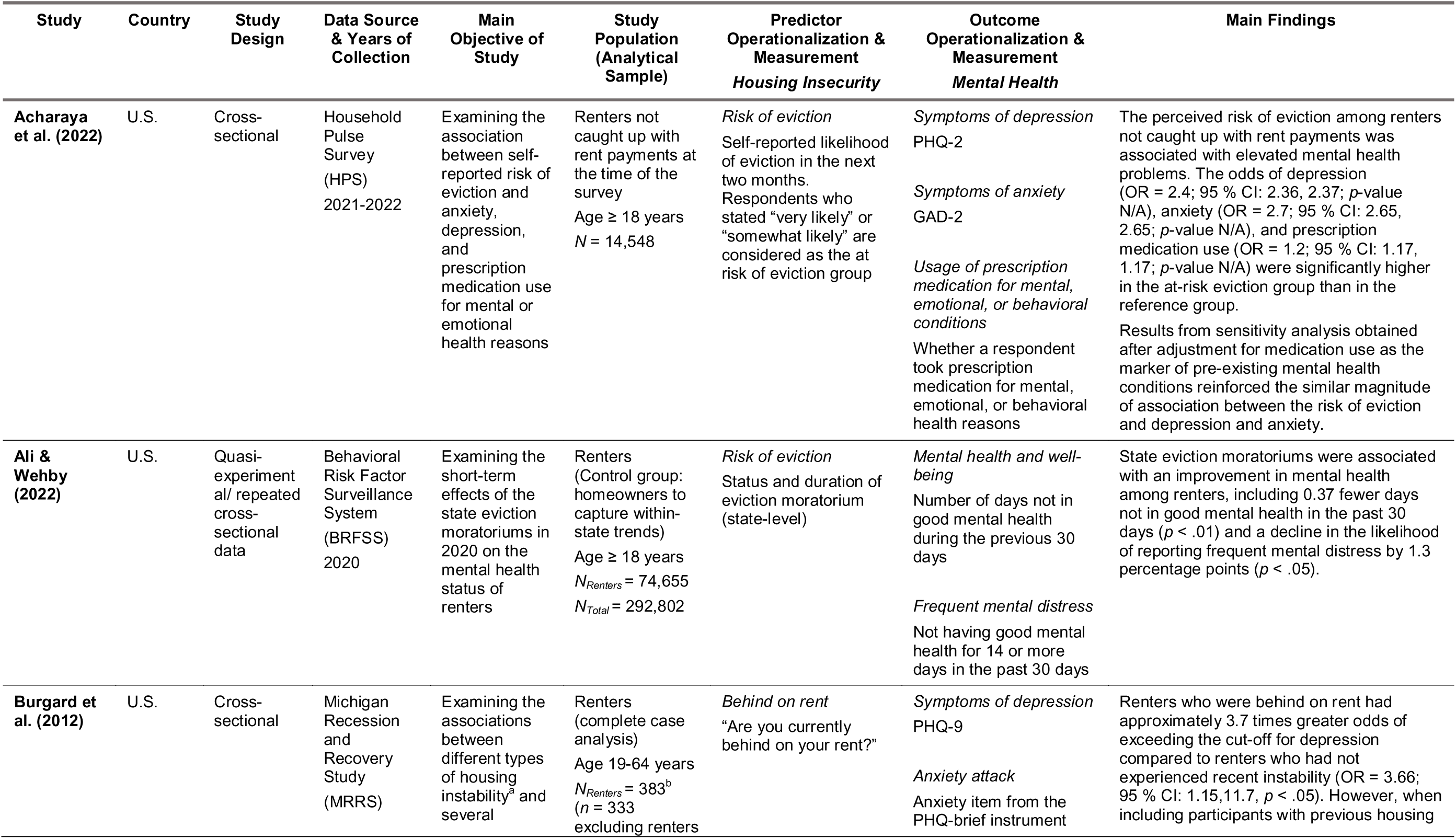
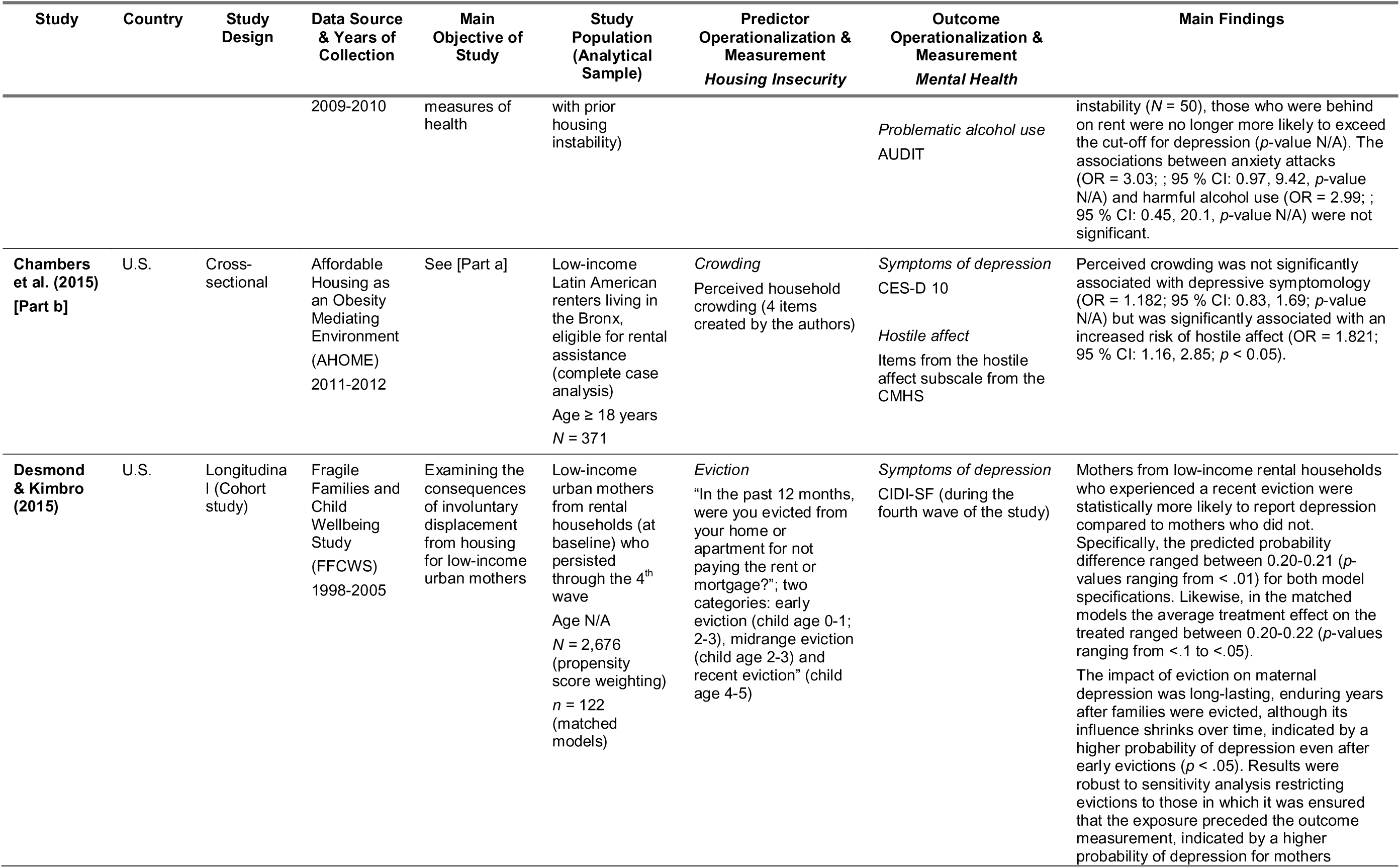
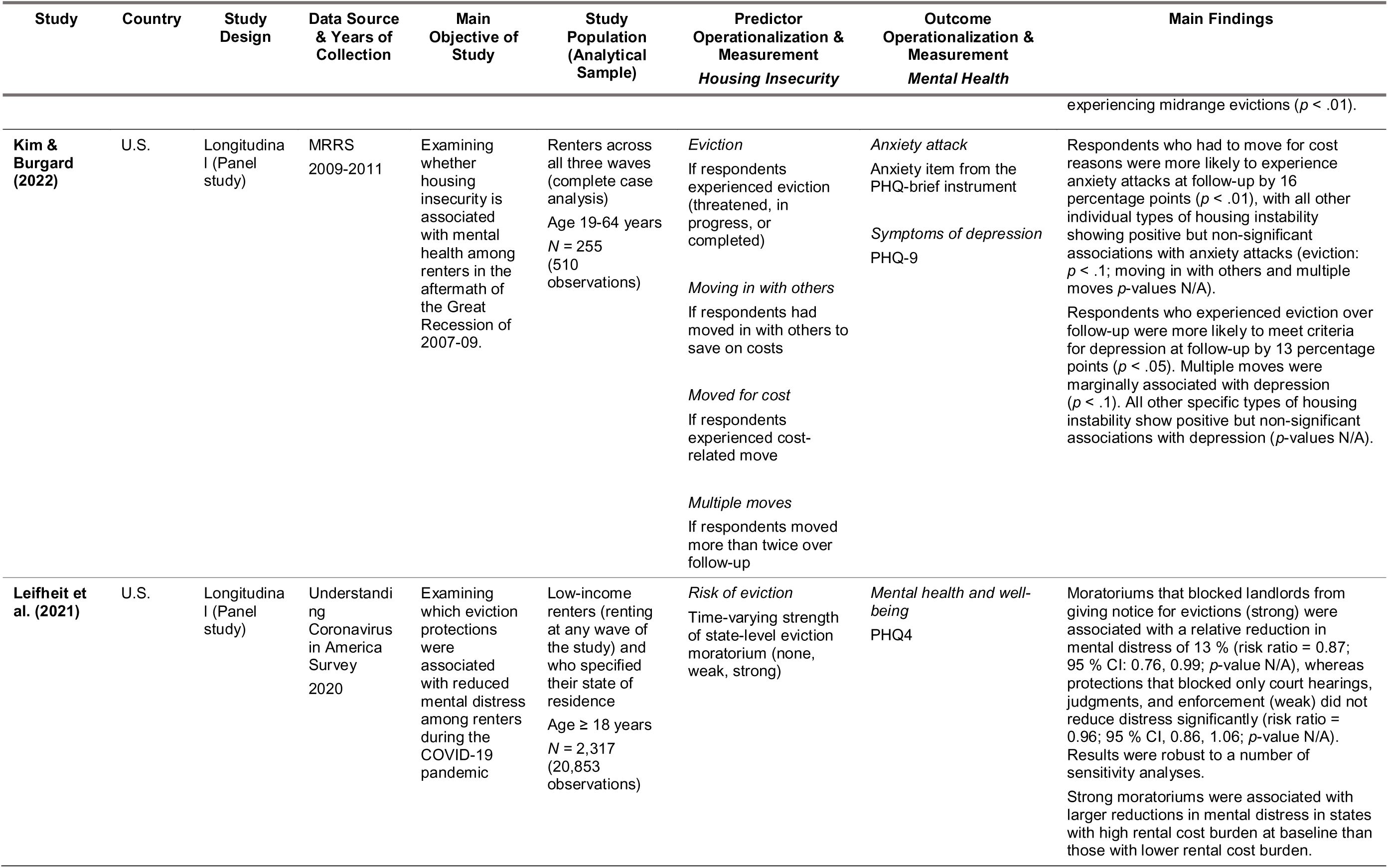
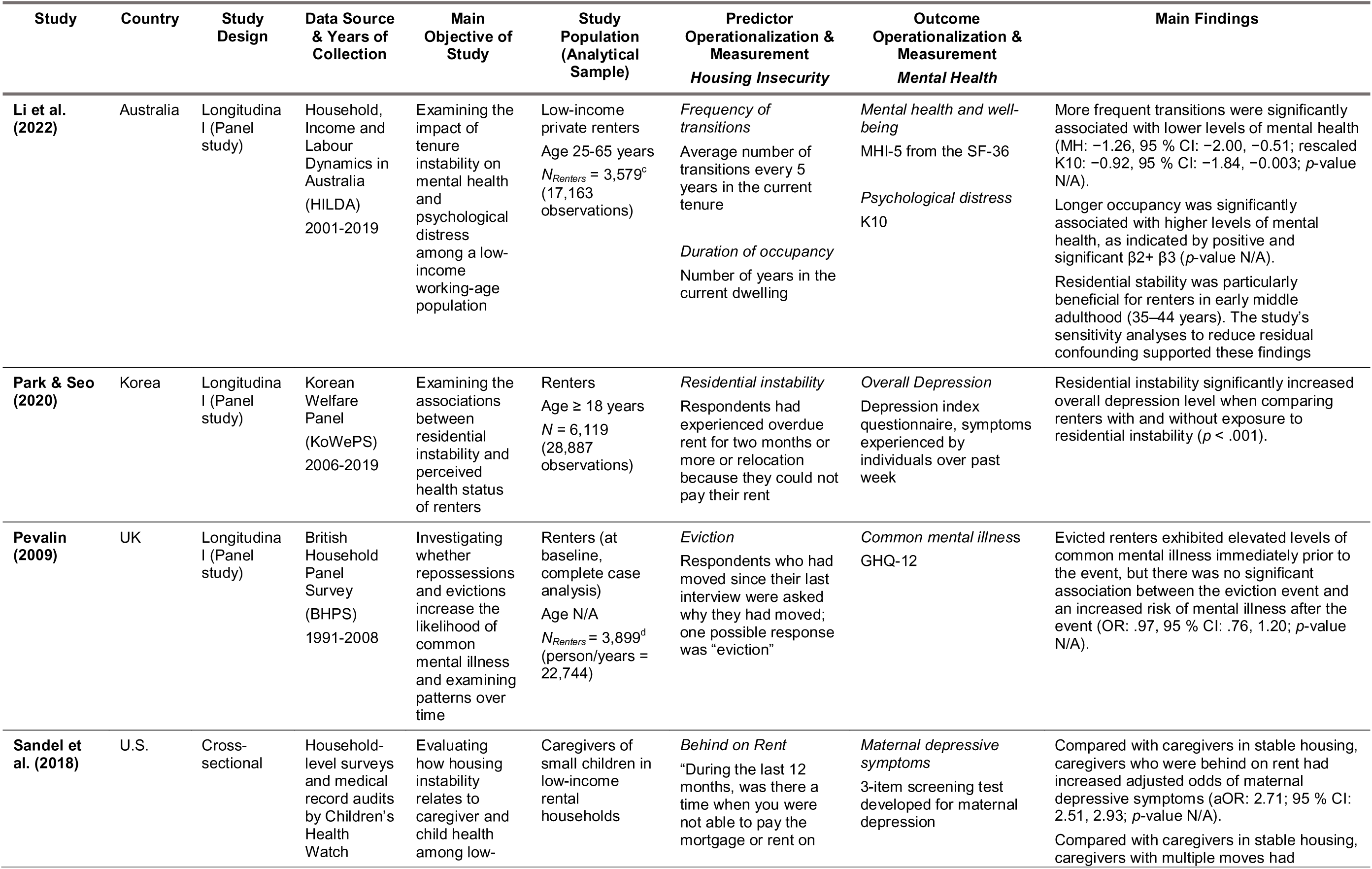
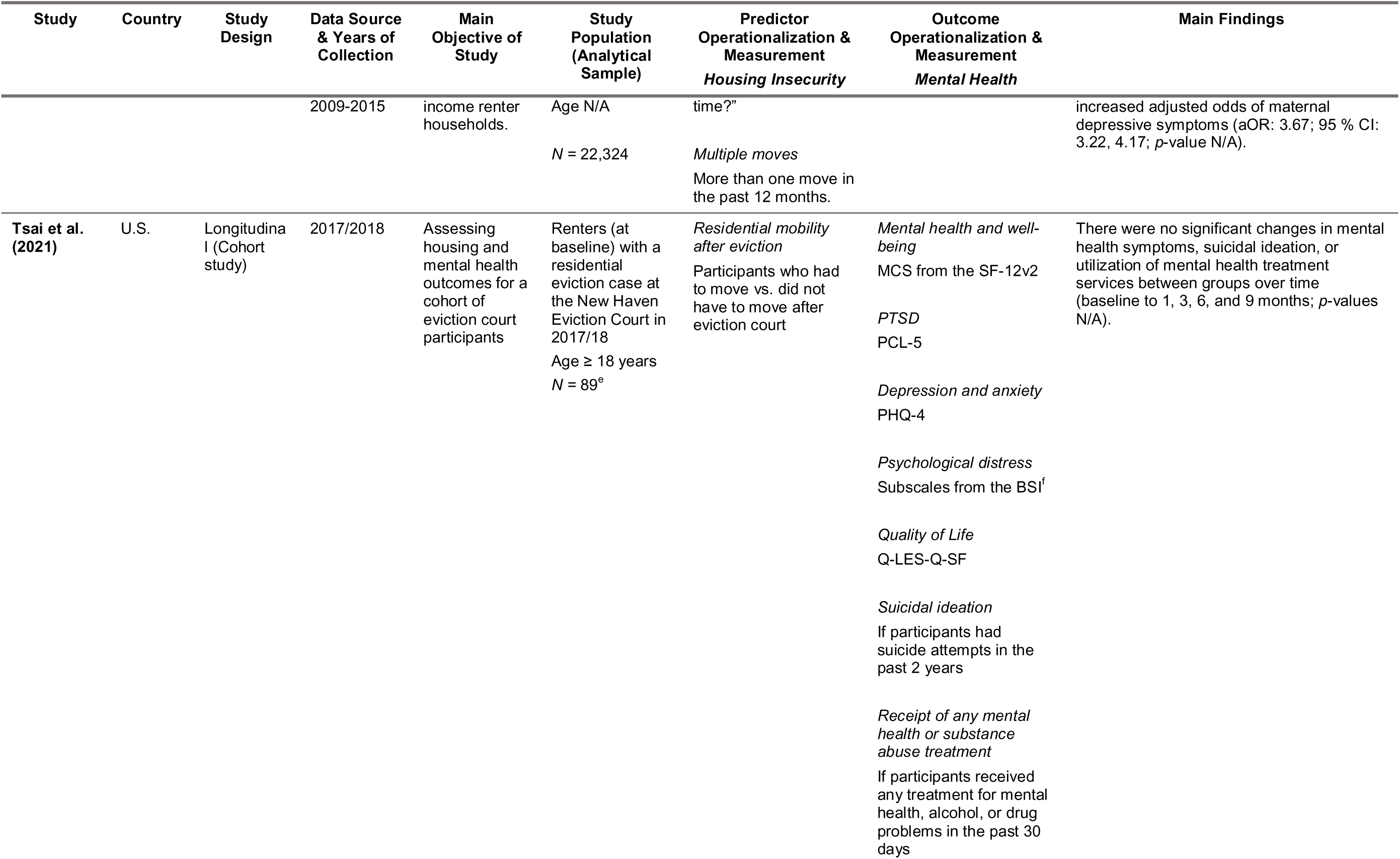
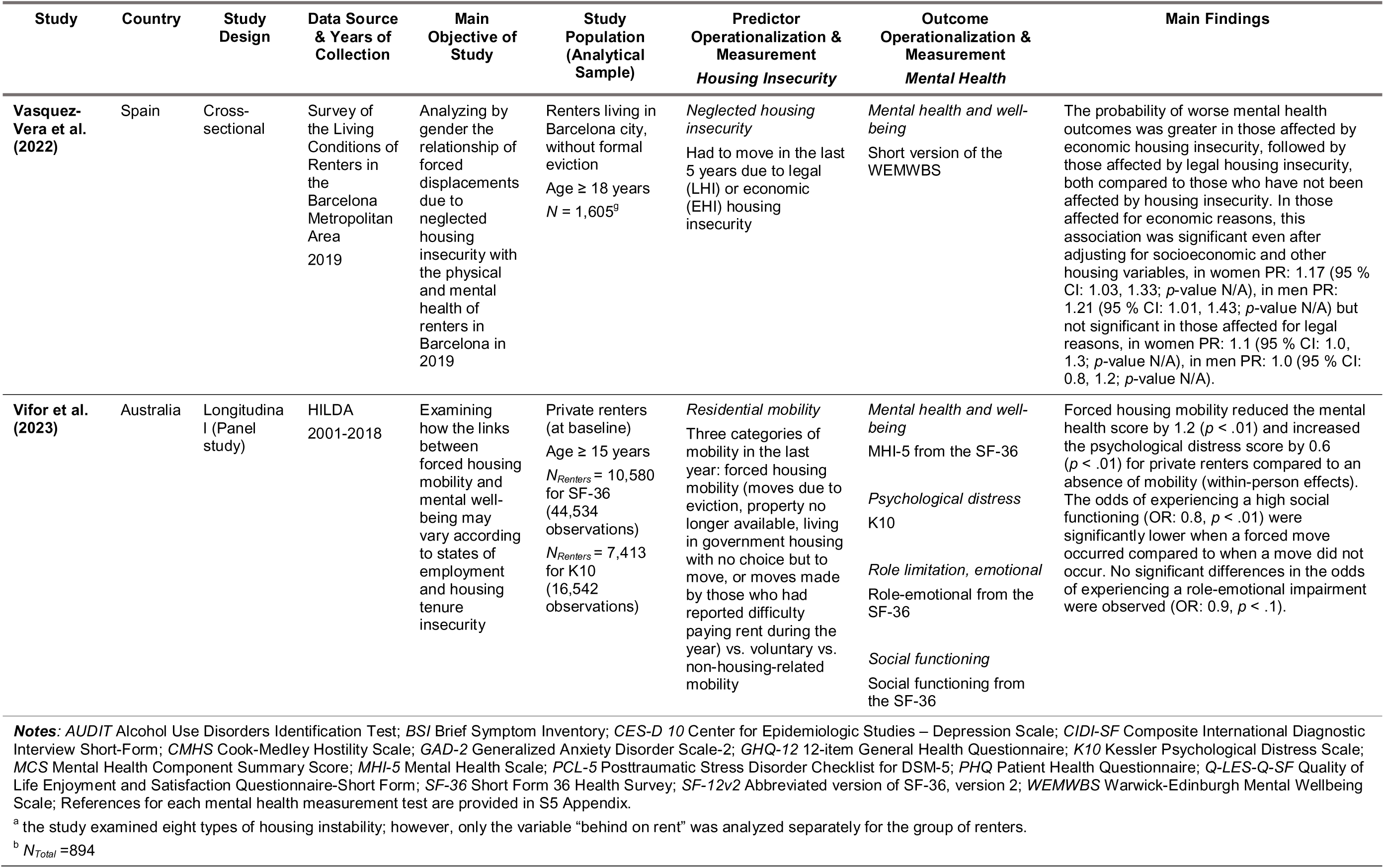
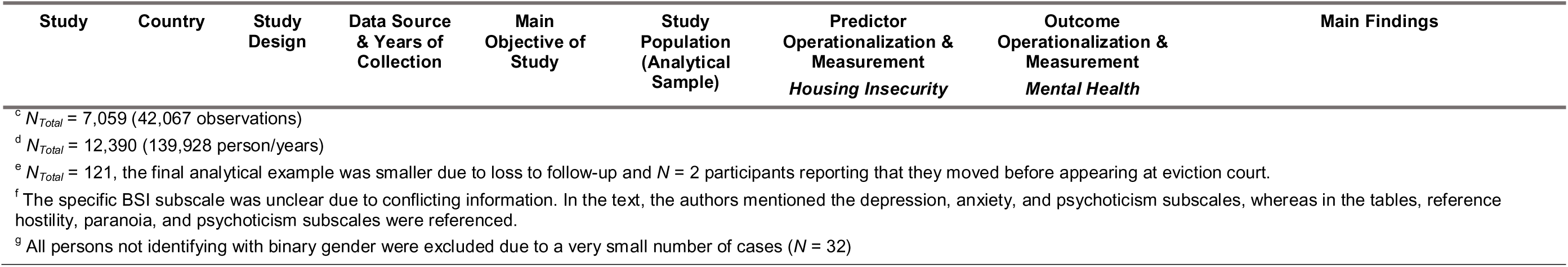
Summary characteristics of the included studies regarding housing instability and mental health (ordered alphabetically according to author’s name)

The selected studies addressed a wide range of research questions, with only a few of them focusing exclusively on the population of interest. Eight out of 22 studies examined housing insecurity broadly, including not only renters but also other housing tenures such as homeowners, while analyzing renters separately in at least one analysis. Six studies identified their (sub-)samples as private renters (Baker, Lester, et al., 2020; Bentley et al., 2016; A. Li et al., 2022; Mason et al., 2013; Reeves et al., 2016; ViforJ et al., 2022) and two as public renters (Baker, Lester, et al., 2020; Prentice & Scutella, 2020). Moreover, a number of studies examined specific population groups, such as low-income renter households (for more details see Table 1 and Table 2. It is important to note that four studies with longitudinal data only measured tenure status at one time point; therefore, it is possible that changes in tenure status occurred over time (Pevalin, 2009; Prentice & Scutella, 2020; J. Tsai et al., 2021; ViforJ et al., 2022).

### Exposure measurements of housing insecurity

Of the 22 studies included in the review, eight focused solely on housing affordability, while 13 examined housing instability (see Table 3), and one study investigated both dimensions. Housing affordability was primarily operationalized using some form of rent-to-income ratio, while housing instability was mainly assessed through indicators of forced moves. With the exception of three studies that employed population-level aggregated measures (Ali & Wehby, 2022; Leifheit et al., 2021; Rodgers et al., 2019), the remaining studies relied on individual participant and self-reported information to assess housing insecurity.

**Table 3.**
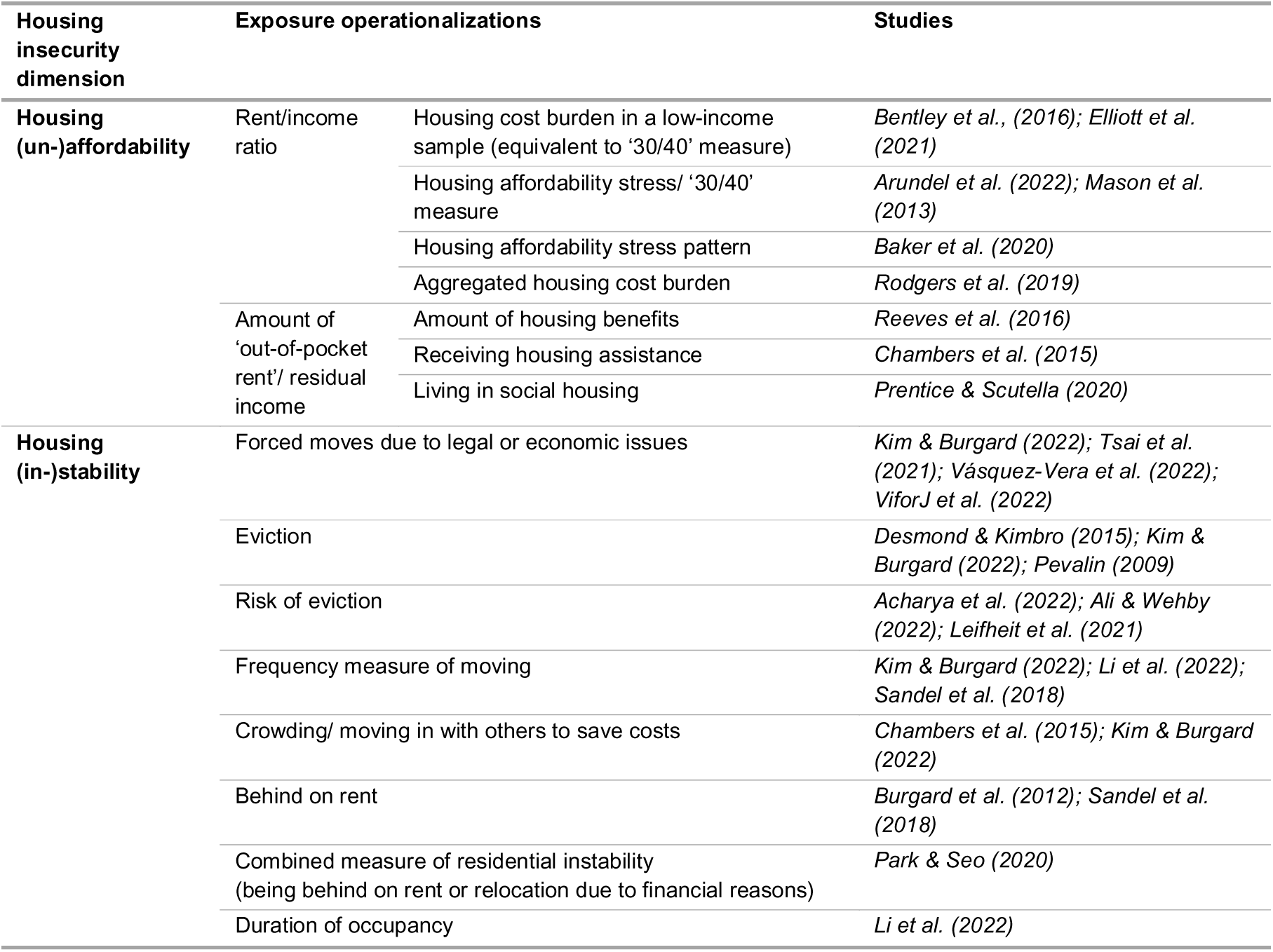
Summary of exposure operationalizations used in the included studies.

### Outcome measurement of mental health

The studies included in the review employed heterogeneous operationalizations of mental health outcomes (see Table 4). Twelve studies measured overall mental health, well-being, and/or distress, summarized here as “overall mental health”. Three studies used measures of psychosocial functioning, such as hostility or interpersonal functioning. Additionally, 11 studies focused on symptoms of mental disorders, primarily examining symptoms of depression. One study investigated suicidal ideation. Two studies explored changes in mental health treatment, including the prescription of medication for mental health purposes. The majority of the selected studies utilized screening tools to measure symptoms of mental disorders, such as the Patient Health Questionnaire-2 (PHQ-2; Kroenke et al., 2003) or screening for maternal depression. Moreover, with the exception of one study in which trained clinical interviewers conducted home interviews (Chambers et al., 2015) and two studies that used a clinical-diagnostic interview (Desmond & Kimbro, 2015; Elliott et al., 2021), all other studies relied on self-report outcome instruments.

**Table 4.**
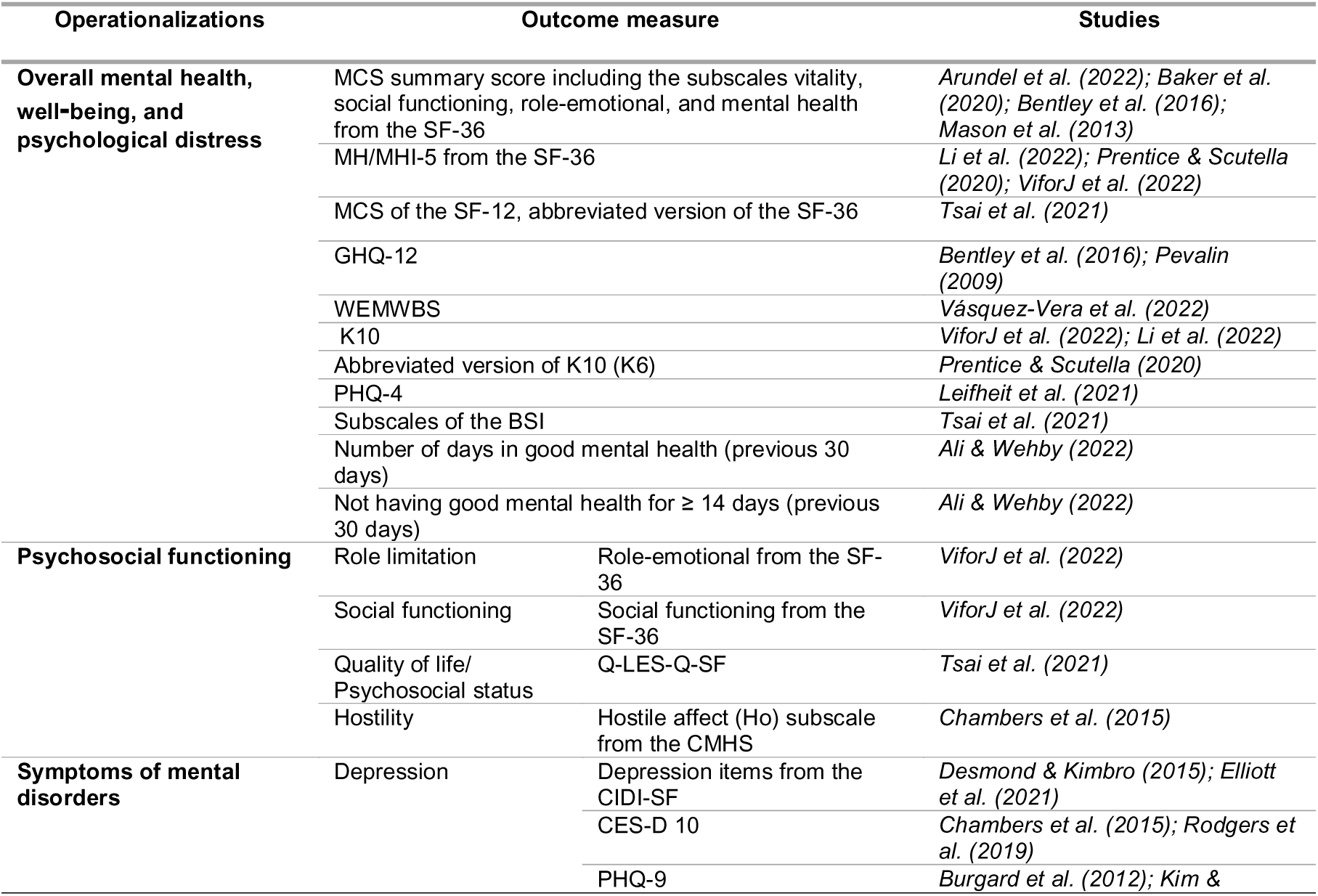
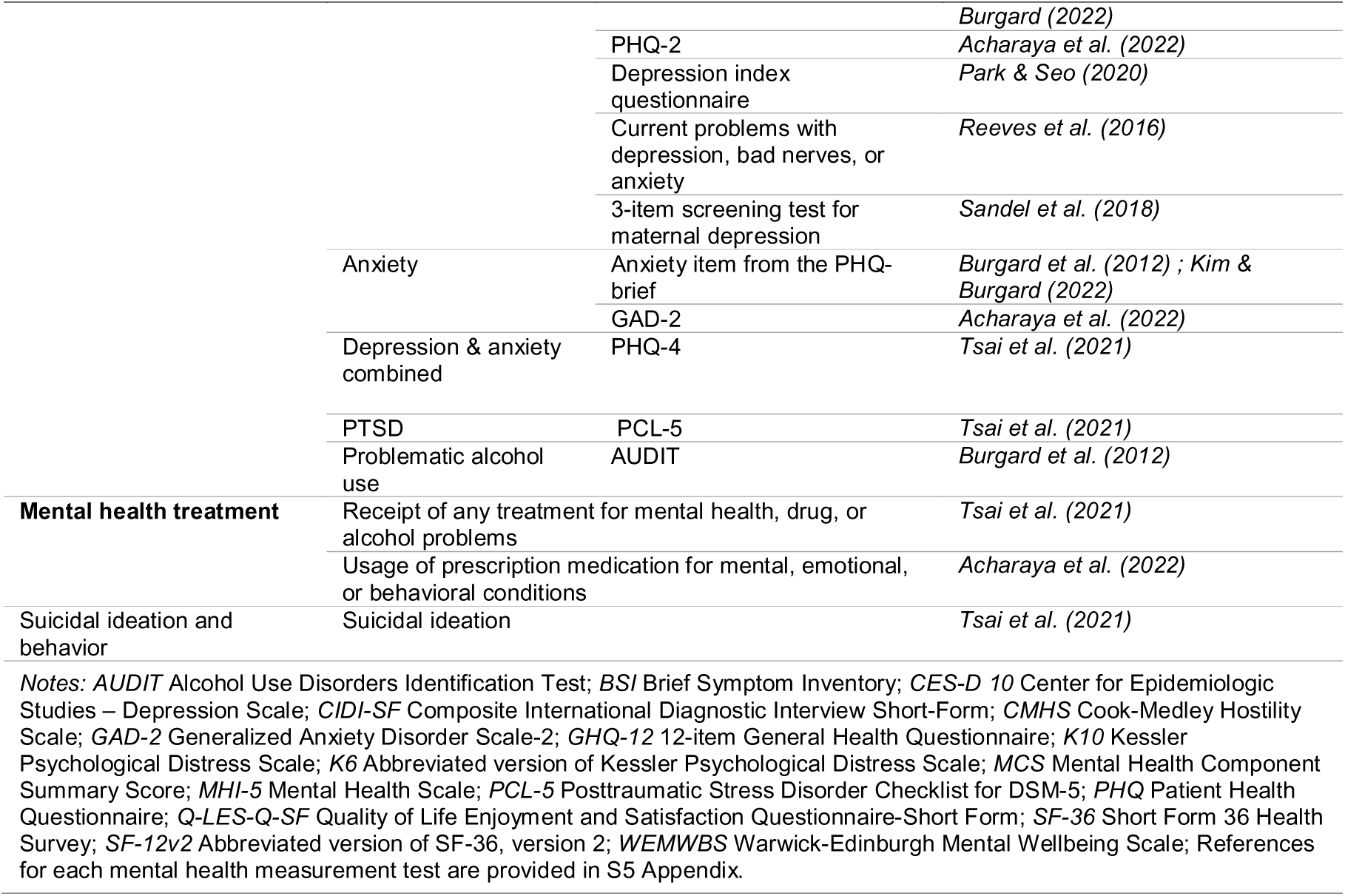
Summary of outcome operationalizations and measurement instruments used in the included studies.

### Reporting quality

A detailed evaluation of every item of the respective reporting guidelines for each study is provided in S4 Appendix. With the exception of Leifheit et al. (2021), none of the included studies explicitly stated adherence to reporting guidelines. Despite this, the included studies generally demonstrated satisfactory reporting quality, with authors transparently reporting essential information such as rationale and objectives, employed variables, analytical approaches, descriptive and analytical results, and at least some limitations. Nevertheless, the critical appraisal of the reporting quality revealed some flaws that impeded accurate data extraction, methodological assessment, and potentially the replication of study findings.

One frequently omitted aspect in the method section was the numerical reporting of psychometric properties of the outcome instruments (references to validation studies are not considered sufficient for the STROBE statement). Additionally, some studies did not address the psychometric properties of the measurements of housing insecurity at all. Furthermore, since the majority of studies included in this review were based on secondary analyses of survey data, some specific aspects need to be considered. While all studies reported the source and most important characteristics of the original data set, some did not provide comprehensive information on recruitment, missing data, follow-up procedure, as well as loss to follow-up (for the longitudinal studies), study size, and funding source. Regarding the findings, a number of studies did not report confidence intervals and most studies did not report unadjusted results. Moreover, an evaluation using the TREND statement indicated specific omissions in the studies using quasi-experimental methods. These include missing details about the “intervention” (item 4), such as content and exposure time span, as well as missing baseline characteristics and comparison of the study groups at baseline (item 14).

The studies employed a variety of statistical models based on their individual study design and dataset. Most of the studies utilized appropriate and robust methods, taking into account relevant covariates. However, two studies conducted only bivariate analyses without accounting for potential confounders, which indicates a significant risk of bias (Baker, Lester, et al., 2020; J. Tsai et al., 2021). Additionally, three studies (Chambers et al., 2015; Sandel et al., 2018; C. Vásquez-Vera et al., 2022) failed to consider income as an important potential confounder.

### Study results

#### Housing Affordability and Mental Health

Of the nine studies examined in this review that investigated the association between housing affordability and mental health among renters, six found that unaffordable housing was significantly associated with a decline in mental health. Five of these studies employed a longitudinal design (Arundel et al., 2022; Baker, Lester, et al., 2020; Bentley et al., 2016; Elliott et al., 2021; Mason et al., 2013) and one a quasi-experimental design (Reeves et al., 2016). One quasi-experimental study (Prentice & Scutella, 2020) reported an inverse association, while one cross-sectional (Chambers et al., 2015) and one longitudinal study (Rodgers et al., 2019) found no significant association between housing affordability and mental health.

The majority of the studies (eight out of nine) on affordability focused on the low-income population, either directly, by limiting their sample to low-income households, or indirectly, by utilizing the ‘30/40’ measure. Only one study, conducted by Rodgers et al. (2019), analyzed the general population of renters, while another study, by Mason et al. (2013), conducted an additional analysis investigating high-income renters; both studies found no association. Among the findings specific to low-income renters, six showed statistical significance (Arundel et al., 2022; Baker et al., 2019; Baker, Lester, et al., 2020; Elliott et al., 2021; Mason et al., 2013; Reeves et al., 2016), whereas two yielded non-significant results (Chambers et al., 2015; Prentice & Scutella, 2020).

When considering only those studies that measured overall mental health, the majority (four out of five) demonstrated a significant association between unaffordable housing and worse overall mental health. Specifically, three studies found that high rental burden had a negative impact on the overall mental health of both Australian public renters (Baker, Lester, et al., 2020) and private renters (Bentley et al., 2016; Mason et al., 2013). A similar trend was confirmed by Arundel et al. (2022) for the Netherlands. Two of the studies reported no significant association for private renters in the UK (Bentley et al., 2016) and in Australia (Baker, Lester, et al., 2020). Prentice and Scutella (2020) found an inverse association, indicating that those living in social housing had significantly worse overall mental health than the matched control group in the private rental sector.

Four studies explored the association between housing affordability and depressive symptoms among renters. Elliott et al. (2021) found a significant positive association between rent burden and maternal depression. Rodgers et al. (2019) found a positive association between aggregated rent burden and depressive symptoms in a sample across tenures, which was not statistically significant when only the smaller subsample of renters was analyzed. Reeves et al. (2016) observed a significant increase in renters’ symptoms of depression after the reduction of housing benefits. In a sample of low-income Latin American renters, neither depressive symptoms nor hostile affect were significantly associated with receiving rental assistance (Chambers et al., 2015).

#### Housing Instability and Mental Health

Of the 14 studies that examined the link between housing instability and mental health, 12 reported one or more significant associations between unstable housing and poorer mental health among renters. Six of these studies employed a longitudinal design (Desmond & Kimbro, 2015; Kim & Burgard, 2022; Leifheit et al., 2021; A. Li et al., 2022; Park & Seo, 2020; ViforJ et al., 2022), five a cross-sectional design (Acharya et al., 2022; Burgard et al., 2012; Chambers et al., 2015; Sandel et al., 2018; C. Vásquez-Vera et al., 2022), and one a quasi-experimental design (Ali & Wehby, 2022). Two longitudinal studies found no significant associations (Pevalin, 2009; J. Tsai et al., 2021).

When considering only those studies that specifically measured overall mental health, five out of seven reported a significant association between unstable housing and poorer mental health. Specifically, Leifheit et al. (2021) and Ali and Wehby (2022) observed a positive association between the eviction moratorium implemented in the U.S. in response to the COVID-19 pandemic and improved mental health of renters. Additionally, three studies found that experiencing forced mobility (C. Vásquez-Vera et al., 2022; ViforJ et al., 2022), or a higher frequency of moves (A. Li et al., 2022), respectively, were associated with a negative impact on the mental health of private renters in Australia and Spain. Conversely, longer duration in the rental property was linked to improved mental health (A. Li et al., 2022). Two studies which focused on eviction (Pevalin, 2009) and forced mobility after eviction (J. Tsai et al., 2021) did not find any statistically significant association with overall mental health.

When examining the studies that investigated depressive symptoms as outcome measures, the majority (seven out of eight) found a significant association between unstable housing and a rise in depressive symptoms. Indicators including (risk of) eviction, forced moves, multiple moves, and being behind on rent showed a link to increased depressive symptoms (Acharya et al., 2022; Burgard et al., 2012; Chambers et al., 2015; Desmond & Kimbro, 2015; Kim & Burgard, 2022; Park & Seo, 2020; Sandel et al., 2018). However, two studies, which examined subjective crowding (Chambers et al., 2015) and moving in with others as well as forced mobility (Kim & Burgard, 2022), did not find a significant association with depressive symptoms.

Three studies examining symptoms of anxiety were included in the review. The results revealed that subjective risk of eviction (Acharya et al., 2022) and forced mobility (Kim & Burgard, 2022) were significantly associated with greater anxiety symptoms. The other measures examined in the study by Kim and Burgard (2022), including eviction, multiple moves, and moving in with others, as well as being behind on rent in the study by Burgard et al. (2012), were not found to be significantly associated with anxiety.

Furthermore, the review included one study on symptoms of PTSD (J. Tsai et al., 2021), one on alcohol abuse (Burgard et al., 2012), and one on a combined measure of depression and anxiety (J. Tsai et al., 2021). J. Tsai et al. (2021) was the only study to investigate suicidal ideation. None of these studies found a significant association between unstable housing and these mental health outcomes. However, one study reported a higher prevalence of prescription medication for mental health reasons (Acharya et al., 2022), while mental health treatment as an outcome measure was not found to be significant in the other study (J. Tsai et al., 2021). Finally, two studies reported a significant association between unstable housing and impairments in social and interpersonal functioning (Chambers et al., 2015; ViforJ et al., 2022).

## Discussion

### Main findings

This systematic review aimed to examine the association between housing insecurity and mental health among renters, with a specific focus on exposure to unaffordability and instability as key dimensions of housing. Through a comprehensive search process, a total of 22 studies that met the inclusion criteria were included.

#### Housing Affordability

Among the nine studies examining housing affordability, six reported significant associations between unaffordable rent, more specifically rent burden and reduction in housing benefits, and low mental health. The majority of these studies (seven out of nine) focused solely on the low-income renter population. Thus, we can conclude from this review an association between unaffordable rent and poor mental health for this population. Notably, the studies with significant findings all took income into account, mitigating the risk of conflating the effect of (low) income and unaffordable rent on mental health (Baker, Lester, et al., 2020). In addition, previous publications also suggest that the burden of unaffordable housing mainly affects low-income renter and homeowner households, whereas high-income households do not experience such effects (Bentley et al., 2011; Morris et al., 2017). Renters with economic capital are likely to face fewer challenges related to housing insecurity, as they possess various means to compensate for these issues (Morris et al., 2017; Routhier, 2019). Moreover, the synthesis indicates that the association between unaffordable housing and mental health might differ depending on the context, in particular being less pronounced in countries with strong tenure protections. This pattern was observed in studies comparing different countries (Bentley et al., 2016) or exploring the consequences of deterioration of rental policies (Arundel et al., 2022; Reeves et al., 2016). These findings are consistent with other research investigating housing across different countries, although they are not restricted to the population of renters (Acolin, 2022; Herbers & Mulder, 2017; Hulse et al., 2011).

However, in the current review, the two included studies that examined housing affordability interventions (social housing and housing assistance) did not reveal differences in mental health outcomes compared to either no intervention (Chambers et al., 2015) or longitudinally after 6-12 months (Prentice & Scutella, 2020). Recent systematic reviews encompassing households across different tenure statuses have also reported inconsistent evidence regarding the association of interventions promoting affordability and stability with improved mental health outcomes (Chen et al., 2022; Dweik & Woodhall-Melnik, 2022). Therefore, it remains unclear which specific interventions effectively enhance mental health among renters, highlighting the need for further studies in this area.

#### Housing Instability

Among the 14 studies examining housing instability, 12 reported significant associations between unstable housing and poor mental health among renters. The significant associations were established through a diverse range of measures of housing instability, most commonly the risk of eviction, forced mobility, and frequency of transitions. Several studies also demonstrated positive effects of stable rental tenure and state eviction moratoriums in mitigating these adverse impacts. The absence of a standard definition for housing instability is evident from the diverse measures employed in the selected studies. However, the fact that many of these measures demonstrated a significant association with renters’ well-being emphasizes the importance of including a range of exposure measures in a unified definition of housing instability (Cox et al., 2019; Routhier, 2019). This will enable future studies to delve more deeply into the multiple pathways through which housing instability appears to affect renters’ well-being.

### Strength of evidence

The findings of this review contribute to the growing body of research on the social determinants of mental health (Baumgartner et al., 2023; Compton & Shim, 2015; Lund et al., 2018; Marmot & Wilkinson, 2005; Shaw, 2004; Suglia et al., 2015; Swope & Hernández, 2019; WHO & Calouste Gulbenkian Foundation, 2014). The review provides strong evidence to support an association between housing insecurity and impaired mental health, with a substantial number of studies addressing each dimension, i.e. housing affordability and instability. In the case of housing affordability, the available data primarily concerned samples of low-income households, limiting the scope of conclusions to this specific population group. A significant proportion of the studies employed robust study designs, such as longitudinal approaches (*n* = 15). For both housing dimensions, the majority of studies demonstrated significant associations between measures of housing insecurity and overall mental health and depressive symptoms.

In contrast, the evidence for other mental health outcomes such as anxiety, mental health behavior, mental health treatment, suicidal ideation or behavior and psychosocial functioning was more limited and inconsistent. Due to the paucity of studies, further research is necessary to draw definitive conclusions about the associations of these outcomes with housing insecurity. Moreover, most of the outcome measurement tools used in the included studies were screening tools and relied on self-report data, and thus did not provide a clinical diagnosis or a comprehensive assessment of mental disorders, although most have shown good reliability and validity, making them useful for monitoring general mental health (Mawani & Gilmour, 2010). However, two studies relied solely on single-item questions (Ali & Wehby, 2022; Reeves et al., 2016), while another two studies did not provide sufficient detail for an assessment of psychometric properties (Park & Seo, 2020; Sandel et al., 2018).

Despite a noticeable shift in recent years, the number of publications specifically addressing housing insecurity among renters remains limited. Furthermore, the sample sizes were often small due to secondary analyses, leading to low statistical power and hindering stratified analysis. Moreover, the included studies that yielded non-significant findings might also have been underpowered, and an effect may potentially have become detectable with sufficient testing strength (see for example, Rodgers et al., 2019).

Although most findings show associations between unaffordable and unstable housing and mental health in different populations and contexts, based on the current evidence, and due to the methodology of the studies, it is not possible to draw causal conclusions. In particular, it is important to highlight that housing insecurity is intertwined with various other variables that might contribute to mental health problems, including socioeconomic factors, stress and negative life events, neighborhood disadvantages, and other social inequality aspects. In addition, the relationship between housing insecurity and mental health is likely to be bidirectional. Previous research across different tenure statuses suggests that pre-existing mental health conditions increase the risk of housing insecurity outcomes, while housing insecurity also negatively affects mental health (Baker et al., 2013; Marçal, 2021; J. Tsai & Huang, 2019).

### Future research directions

Housing insecurity often manifests across multiple dimensions simultaneously, such as affordability, (in-)stability, and quality, and intersects with further social and economic disadvantages such as gender, disability, and race (Bentley et al., 2019; Carrere et al., 2022; DeLuca & Rosen, 2022; Hulse & Saugeres, 2008; Kirkpatrick & Tarasuk, 2011; Morris et al., 2017). On a micro-level, these factors can accumulate and reinforce each other, amplifying their adverse effects on mental health. Studies further indicate that housing insecurity and its impact on mental health also varies on a macro-level, across different social, legal, and cultural contexts (Acolin, 2022; Bentley et al., 2016; Herbers & Mulder, 2017; Hulse & Milligan, 2014; Hulse et al., 2011). Most of the included studies (19 out of 22) in this review were conducted in the UK, the U.S., and Australia, underlining the need for research from a broader range of countries. This is particularly relevant as housing insecurity is becoming increasingly prevalent, affecting the mental health of renters even in countries with previously good tenure security such as the Netherlands (Arundel et al., 2022). Given the heterogeneity within the group of renters and the likelihood of diverse experiences of housing insecurity and its impact on mental health based on economic and social circumstances, it is therefore crucial that future research adopts an intersectional lens and a bidirectional approach. Future research may additionally incorporate clinical interviews as well as measures that specifically capture other mental health problems potentially linked to housing insecurity, such as anxiety, suicide, and substance abuse (Lund et al., 2018).

### Practical implications

From a clinical perspective, the findings highlight the importance of considering social factors in understanding and effectively treating mental health issues. Health professionals could integrate screening questions for housing insecurity into the assessment, and remain aware of its potential impact throughout treatment, given its potential role as a triggering, maintaining, or reinforcing factor. More generally, adopting community psychological approaches that integrate mental healthcare with social work and legal support might be beneficial. The use of a multidisciplinary approach could include helping tenants who are experiencing housing insecurity to find affordable housing, access housing assistance, or navigate or resolve legal issues. Similar approaches have been successfully employed in homelessness prevention programs that align with the aim of achieving “zero discharge into homelessness” (Gaetz & Dej, 2017, p. 60; Moritsugu et al., 2019). As part of a multidisciplinary approach, health professionals would have a unique opportunity to shape public discourse, drive research efforts, and inform policy discussions to advocate for transformative change in the conditions contributing to social disadvantages (Compton & Shim, 2015). The findings of our review point to the potential benefit of providing affordable and stable housing options for renters, particularly for low-income households, as a strategy to improve public mental health. The question of which precise housing policy measures are effective, particularly for the most vulnerable groups of renters such as low-income, disabled, or racially marginalized households, remains an area for further research and investigation (DeLuca & Rosen, 2022).

### Limitations

It is important to acknowledge some limitations of this systematic review. First, only a random 10 % of the studies underwent double screening, and similarly, data extraction was independently verified for only 30 % of the included studies. This may introduce bias and increase the risk of missed studies. Second, this review focused solely on two dimensions of housing insecurity, i.e. affordability and instability, even though housing insecurity is a multidimensional concept that includes further challenges such as housing conditions, safety, and neighborhood opportunities. Third, we did not specifically explore the mechanisms linking housing insecurity and mental health, which could include factors such as stress, neighborhood conditions, disrupted routines, social integration, social support, stigma, financial stress, and access to healthcare (Compton & Shim, 2015; Kirst et al., 2020; Lund et al., 2018; Suglia et al., 2015; Swope & Hernández, 2019). Fourth, there was considerable heterogeneity between the designs of the individual studies, making it difficult to directly compare the results and to conduct a meta-analysis. Lastly, the eligibility criteria for this review were restricted to OECD countries, limiting the generalizability of the findings. However, some literature from non-OECD countries also points in a similar direction (Acolin, 2022; Clair et al., 2016; Luginaah et al., 2010).

## Conclusion

This systematic review is the first to specifically investigate the association between housing insecurity and mental health among renters, addressing a crucial gap in the literature. The findings indicate an association of renters’ exposure to housing unaffordability and instability with adverse mental health outcomes, including overall mental health and depressive symptoms. Based on the current findings, health professionals might consider housing insecurity as a contributing and aggravating factor regarding mental health issues. Housing insecurity poses a global challenge for renters in OECD countries, highlighting the need for policymakers to implement supportive housing policies and tenure protection measures in order to improve renters’ housing security and ultimately public health.

## Supporting information

S1_Appendix_PRSIMA Checklist

## Data Availability

All relevant data are within the manuscript and its Supporting Information files.

## Acknowledgements

We would like to thank Sarah Mannion for the language editing of the manuscript.

## Supporting information

**S1 Appendix. Preferred Reporting Items for Systematic Reviews and Meta-Analyses (PRISMA) Checklist 2020**.

**S2 Appendix. Complete Search Syntax.**

**S3 Appendix. Screening Form (Full-Text) Including Reasons for Exclusion for Each Study Report.**

**S4 Appendix. Reporting Quality of the Included Studies**.

**S5 Appendix. Reference List of the Measurement Tools Used in the Included Studies**

