## Supplementary material for "Exploring the association between housing insecurity and mental health among renters: A systematic review": S2_Appendix

**S2 Appendix. Complete Search Syntax.**

| **Search components** | **keywords** | | **MEDLINE** | **PsycINFO** | **Web of Science Core Collection**  (Arts & Humanities Citation Index (A&HCI), Book Citation Index - Science (BKCI-S), Book Citation Index - Social Sciences & Humanities (BKCI-SSH), Conference Proceedings Citation Index - Science (CPCI-S), Conference Proceedings Citation Index - Social Science & Humanities (CPCI-SSH), Emerging Sources Citation Index (ESCI), Science Citation Index Expanded (SCI-Expanded), Social Sciences Citation Index (SSCI)) | **Applied Social Sciences Index and Abstracts (ASSIA)** |
| --- | --- | --- | --- | --- | --- | --- |
|  |  |  | **PubMed** | **EBSCO** | **Clarivate Analytics** | **ProQuest** |
|  |  |  | *Title, Abstract, Keywords + MeSH-Terms* | *Title, Abstract, keywords + thesaurus-terms* | *Topic (title, abstract, keywords, keywords+)* | *noft (all excl. full-textt) + theausurs-terms* |
| **Population:**  **Renters** | Rent  Renter  renters  Renting  Rental  rented  Tenant  Tenancy  private rental sector  private rental market  Residents in social housing  Residents in public housing  Rental/ housing assistance  Rental/housing benefit  Rent/housing allowance(s)  Housing/rent/rental subsidy/subsidies  Subsidized/ subsidised Housing | Rent*  Tenan*  “social housing”  “public housing”  “housing assistance”  “housing subsid*”  “housing allowance*”  “housing benefit*”  “subsidi*ed housing” | (rent*[tiab] OR tenan*[tiab] OR "public housing"[tiab] OR "social housing"[tiab] OR "housing assistance"[tiab] OR "housing subsid*"[tiab] OR "housing allowance*"[tiab] OR "housing benefit*"[tiab] OR "subsidised housing"[tiab] "subsidized housing"[tiab]) | ((TI (rent OR renter# OR rented OR renting OR rental OR tenan* OR "public housing" OR “social housing” OR “housing assistance” OR “housing subsid*” OR “housing allowance#” OR “housing benefit#” OR “subsidi?ed housing”)) OR (AB (rent OR renter# OR rented OR renting OR rental OR tenan* OR "public housing" OR “social housing” OR “housing assistance” OR “housing subsid*” OR “housing allowance#” OR “housing benefit#” OR “subsidi?ed housing”)) OR (KW (rent* OR tenan* OR "public housing" OR “social housing” OR “housing assistance” OR “housing subsid*” OR “housing allowance#” OR “housing benefit#” OR “subsidi?ed housing”))) | TS = (rent* OR tenan* OR "public housing" OR “social housing” OR “housing assistance” OR “housing subsid*” OR “housing allowance$” OR “housing benefit$” OR “subsidi$ed housing”) | noft(rent* OR tenan* OR "public housing" OR “social housing” OR "housing assistance" OR "housing subsid*" OR "housing allowance*" OR "housing benefit*" OR "subsidi?ed housing" OR "Housing assistance programmes") |
| **AND** | | | | | | |
| **Risk factor:**  **Housing Insecurity** | Insecurity  Insecure  Secure  Security  Precarity  Housing Crisis  Housing strain  Housing stress | Secur*  Insecur*  Precarity  “housing strain”  “housing stress”  “housing crisis” | (insecur*[tiab] OR secur*[tiab] OR precarity[tiab] OR "housing strain*"[tiab] OR "housing stress"[tiab] OR "housing crisis"[tiab] OR affordab*[tiab] OR unaffordab*[tiab] OR cost*[tiab] OR arrear*[tiab] OR "payment problem*"[tiab] OR "financial strain"[tiab] OR mobility[tiab] OR instab*[tiab] OR stability[tiab] OR stable[tiab] OR unstable[tiab] OR eviction[tiab] OR "multiple moves"[tiab] OR "frequent moves"[tiab] OR "forced moves"[tiab] OR "home loss"[tiab] OR "housing loss"[tiab] OR "duration of stay"[tiab] OR "doubling up"[tiab] OR overcrowding[tiab] OR "Housing Instability"[Mesh]) | ((TI (insecur* OR secur* OR precarity OR “housing strain#” OR “housing stress” OR “housing crisis” OR affordab* OR unaffordab* OR cost# OR arrear# OR “payment problem#” OR “financial strain” OR mobility OR instab* OR stability OR stable OR unstable OR eviction OR "multiple moves" OR "frequent moves" OR "forced moves" OR “home loss” OR "housing loss" OR “duration of stay” OR “doubling up” OR overcrowding)) OR (AB(insecur* OR secur* OR precarity OR “housing strain#” OR “housing stress” OR “housing crisis” OR affordab* OR unaffordab* OR cost# OR arrear# OR “payment problem#” OR “financial strain” OR mobility OR instab* OR stability OR stable OR unstable OR eviction OR "multiple moves" OR "frequent moves" OR "forced moves" OR “home loss” OR "housing loss" OR “duration of stay” OR “doubling up” OR overcrowding)) OR (KW (insecur* OR secur* OR precarity OR “housing strain#” OR “housing stress” OR “housing crisis” OR affordab* OR unaffordab* OR cost# OR arrear# OR “payment problem#” OR mobility OR instab* OR stability OR stable OR unstable OR eviction OR "multiple moves" OR "frequent moves" OR "forced moves" OR “home loss” OR "housing loss" OR “duration of stay” OR “doubling up” OR overcrowding))) | TS = (insecur* OR secur* OR precarity OR “housing strain$” OR “housing stress” OR “housing crisis” OR affordab* OR unaffordab* OR cost$ OR arrear$ OR “payment problem$” OR “financial strain” OR mobility OR instab* OR stability OR stable OR unstable OR eviction OR "multiple moves" OR "frequent moves" OR "forced moves" OR “home loss” OR "housing loss" OR “duration of stay” OR “doubling up” OR overcrowding) | noft(insecur* OR secur* OR precarity OR "housing strain*" OR "housing stress" OR "housing crisis" OR affordab* OR unaffordab* OR cost* OR arrear* OR "payment problem*" OR mobility OR Instab* OR stability OR stable OR unstable OR eviction OR "multiple moves" OR "frequent moves" OR "forced moves" OR "home loss" OR "housing loss" OR "duration of stay" OR "doubling up" OR overcrowding) |
|  | Affordability  Affordable  Unaffordable  Unaffordability  Cost  Costs  Cost burden  Housing affordability stress  housing payment problems  housing/rent arrears  financial strain | Affordab*  Unaffordab*  Cost*  “payment problems”  Arrear*  “financial strain” |  |  |  |  |
|  | Residential/housing mobility  Instability  Unstable  Stability  Stable  Forced moves  Frequent moves  Multiple moves  Eviction  Home loss  Housing loss  Duration of stay  doubling up  overcrowding | Mobility  Instability  Unstable  Stability  Stable  “Forced moves”  “Frequent moves”  “Multiple moves”  Eviction  “home loss”  “housing loss”  “duration of stay”  “doubling up”  overcrowding |  |  |  |  |
| **AND** | | | | | | |
| **Outcome:**  **Mental Health** | Mental health  Mental Disorder  Mental well-being  Mental stress  Mental illness | Mental | (mental[tiab] OR psych*[tiab] OR "well?being"[tiab] OR depress*[tiab] OR "mood disorder*"[tiab] OR "affective disorder*"[tiab] OR "bipolar disorder*"[tiab] OR Anxiety[tiab] OR "neurotic disorder*"[tiab] OR schizo*[tiab] OR "delusional disorder*"[tiab] OR "somatoform disorder*"[tiab] OR "stress?related disorder*"[tiab] OR "stress disorder*"[tiab] OR "Obsessive?compulsive"[tiab] OR "trauma disorder*"[tiab] OR "dissociative disorder*"[tiab] OR "somatic symptom disorder*"[tiab] OR "eating disorder*"[tiab] OR "sleep?wake disorder*"[tiab] OR "sexual dysfunction"[tiab] OR "paraphilic disorder*"[tiab] OR "personality disorder*"[tiab] OR "gender dysphoria"[tiab] OR "disruptive disorder*"[tiab] OR "impulse?control disorder*"[tiab] OR "conduct disorder*"[tiab] OR "behaviour* disorder*"[tiab] OR "behavior* disorder*"[tiab] OR "substance?related disorder*"[tiab] OR "addictive disorder*"[tiab] OR "substance use"[tiab] OR "neurocognitive disorder*"[tiab] OR "emotional distress"[tiab] OR "emotional disorder"[tiab] OR suicid*[tiab] OR "Mental Disorders"[Mesh] OR "Mental Health"[Mesh] OR "Stress, Psychological"[Mesh] OR "Psychological Distress"[Mesh] OR "Suicide"[Mesh]) | ((TI (mental OR psych* OR "well#being" OR depress* OR “mood disorder#” OR “affective disorder” OR “bipolar disorder#” OR “emotional disorder” OR Anxiety OR “neurotic disorder#” OR schizo* OR “delusional disorder#” OR “somatoform disorder#” OR “stress#related disorder#” OR “stress disorder#” OR “Obsessive#compulsive” OR “trauma disorder#” OR “dissociative disorder#” OR “somatic symptom disorder#” OR “eating disorder#” OR “sleep#wake disorder#” OR “sexual dysfunction#” OR “paraphilic disorder#” OR “personality disorder#” OR “gender dysphoria” OR “disruptive disorder#” OR “impulse#control disorder#” OR “conduct disorder#” OR “behavio#r* disorder#” OR “substance#related disorder#” OR “addictive disorder#” OR “substance use” OR “neurocognitive disorder#” OR suicid*)) OR (AB (mental OR psych* OR "well#being" OR depress* OR “mood disorder#” OR “affective disorder” OR “bipolar disorder#” OR “emotional disorder” OR Anxiety OR “neurotic disorder#” OR schizophrenia OR schizotypal OR “delusional disorder#” OR “somatoform disorder#” OR “stress#related disorder#” OR “stress disorder#” OR “Obsessive#compulsive” OR “trauma disorder#” OR “dissociative disorder#” OR “somatic symptom disorder#” OR “eating disorder#” OR “sleep#wake disorder#” OR “sexual dysfunction#” OR “paraphilic disorder#” OR “personality disorder#” OR “gender dysphoria” OR “disruptive disorder#” OR “impulse#control disorder#” OR “conduct disorder#” OR “behavio#r* disorder#”OR “substance#related disorder#” OR “addictive disorder#” OR “substance use” OR “neurocognitive disorder#” OR suicid*)) OR (DE ("Well Being" OR "Subjective Well Being" OR "Mental Health" OR "Emotional Health" OR "Psychological Stress" OR "Psychiatric Symptoms" OR "Psychological Consequence" OR "Psychological Stress" OR "Psychopathology" OR "Psychosocial Factors" OR "Psychosocial Outcomes" OR "Mental Disorders" OR "Affective Disorders" OR "Disruptive Mood Dysregulation Disorder" OR "Major Depression" OR "Seasonal Affective Disorder" OR "Anxiety Disorders" OR "Castration Anxiety" OR "Generalized Anxiety Disorder" OR "Obsessive Compulsive Disorder" OR "Panic Attack" OR "Panic Disorder" OR "Phobias" OR "Separation Anxiety Disorder" OR "Trichotillomania" OR "Bipolar Disorder" OR "Borderline States" OR "Chronic Mental Illness" OR "Dissociative Disorders" OR "Eating Disorders" OR "Gender Dysphoria" OR "Mental Disorders due to General Medical Conditions" OR "Neurocognitive Disorders" OR "Neurodevelopmental Disorders" OR "Neurosis" OR "Paraphilias" OR "Personality Disorders" OR "Psychosis" OR "Serious Mental Illness" OR "Sleep Wake Disorders" OR "Somatoform Disorders" OR "Stress and Trauma Related Disorders" OR "Substance Related and Addictive Disorders" OR "Thought Disorders" OR "Anxiety" OR "Depression (Emotion)" OR "Suicidal Behavior" OR "Attempted Suicide" OR "Suicidal Ideation" OR "Suicide"))) | TS = (mental OR psych* OR "well$being" OR depress* OR “mood disorder$” OR “affective disorder” OR “bipolar disorder$” OR “emotional disorder$” OR Anxiety OR “neurotic disorder$” OR schizo* OR “delusional disorder$” OR “somatoform disorder$” OR “stress*related disorder$” OR “stress disorder$” OR “obsessive$compulsive” OR “trauma disorder$” OR “dissociative disorder$” OR “somatic symptom disorder$” OR “eating disorder$” OR “sleep*wake disorder$” OR “sexual dysfunction$” OR “paraphilic disorder$” OR “personality disorder$” OR “gender dysphoria” OR “disruptive disorder$” OR “impulse*control disorder$” OR “conduct disorder$” OR “behavi$r* disorder” OR “substance*related disorder$” OR “addictive disorder$” OR “substance use” OR “neurocognitive disorder$” OR suicid*) | noft(mental OR psych* OR "well*being" OR depress* OR "mood disorder*" OR "affective disorder" OR "bipolar disorder*" OR “emotional disorder” OR Anxiety OR "neurotic disorder*" OR schizo* OR "delusional disorder*" OR "somatoform disorder*" OR "stress*related disorder*" OR "stress disorder*" OR "Obsessive*compulsive" OR "trauma disorder*" OR "dissociative disorder*" OR "somatic symptom disorder*" OR "eating disorder*" OR "sleep*wake disorder*" OR "sexual dysfunction*" OR "paraphilic disorder*" OR "personality disorder*" OR "gender dysphoria" OR "disruptive disorder*" OR "impulse*control disorder*" OR "conduct disorder*" OR “behavio*r disorder*” OR "substance*related disorder*" OR "addictive disorder*" OR "substance use" OR "neurocognitive disorder*" OR “emotional distress” OR suicid*) |
|  | Psychological distress  Psychological health  Psychological stress  Psychological illness  Psychological Well-being  Psychological morbidity  Psychological mortality  Psychological symptoms  Psychosocial  Psycho-social | Psych* |  |  |  |  |
|  | Psychiatric  Psychiatrical  Psychiatrical health  Psychiatric illness  Psychiatric disorder  Psychiatric symptoms  Psychiatrical morbidity  Psychiatrical mortality |  |  |  |  |  |
|  | Well-being  Wellbeing  Well being  Emotional disorder  Emotional well-being | “well*being”  Emotion* |  |  |  |  |
|  | Suicide  Suicidal  suicidality  Suicidal behaviour/ideation  Attempted suicide | Suicid* |  |  |  |  |
|  | Specific mental disorders (categories from DSM-5 and ICD-10):  Schizophrenia, psychotic, psychosis, schizotypal, delusional disorder(s), Depression, depressive, mood disorder(s), affective disorder(s), affective symptom(s), bipolar disorder(s), Anxiety, neurotic disorder(s), somatoform disorder(s), stress-related disorder(s), stress disorder(s), Obsessive-compulsive, trauma disorder(s), dissociative disorder(s), somatic symptom disorder(s), eating disorder(s), sleep-wake disorder(s), sexual dysfunction, paraphilic disorder(s), personality disorder(s), gender dysphoria, disruptive disorder(s), impulse-control disorder(s), conduct disorder(s), behavio(u)r(al) disorder(s), substance-related disorder(s), addictive disorder(s), neurocognitive disorder(s) | depress* OR "mood disorder*" OR "affective disorder" OR "bipolar disorder*" OR “emotional disorder” OR Anxiety OR "neurotic disorder*" OR schizo* OR "delusional disorder*" OR "somatoform disorder*" OR "stress*related disorder*" OR "stress disorder*" OR "Obsessive*compulsive" OR "trauma disorder*" OR "dissociative disorder*" OR "somatic symptom disorder*" OR "eating disorder*" OR "sleep*wake disorder*" OR "sexual dysfunction*" OR "paraphilic disorder*" OR "personality disorder*" OR "gender dysphoria" OR "disruptive disorder*" OR "impulse*control disorder*" OR "conduct disorder*" OR “behavio*r disorder*” OR "substance*related disorder*" OR "addictive disorder*" OR "substance use" OR "neurocognitive disorder*" |  |  |  |  |
| **Final syntax** | | | (rent*[tiab] OR tenan*[tiab] OR "public housing"[tiab] OR "social housing"[tiab] OR "housing assistance"[tiab] OR "housing subsid*"[tiab] OR "housing allowance*"[tiab] OR "housing benefit*"[tiab] OR "subsidized housing"[tiab] OR "subsidised housing"[tiab]) AND (mental[tiab] OR psych*[tiab] OR "well?being"[tiab] OR depress*[tiab] OR "mood disorder*"[tiab] OR "affective disorder*"[tiab] OR "bipolar disorder*"[tiab] OR Anxiety[tiab] OR "neurotic disorder*"[tiab] OR schizo*[tiab] OR "delusional disorder*"[tiab] OR "somatoform disorder*"[tiab] OR "stress?related disorder*"[tiab] OR "stress disorder*"[tiab] OR "Obsessive?compulsive"[tiab] OR "trauma disorder*"[tiab] OR "dissociative disorder*"[tiab] OR "somatic symptom disorder*"[tiab] OR "eating disorder*"[tiab] OR "sleep?wake disorder*"[tiab] OR "sexual dysfunction"[tiab] OR "paraphilic disorder*"[tiab] OR "personality disorder*"[tiab] OR "gender dysphoria"[tiab] OR "disruptive disorder*"[tiab] OR "impulse?control disorder*"[tiab] OR "conduct disorder*"[tiab] OR "behaviour* disorder*"[tiab] OR "behavior* disorder*"[tiab] OR "substance?related disorder*"[tiab] OR "addictive disorder*"[tiab] OR "substance use"[tiab] OR "neurocognitive disorder*"[tiab] OR "emotional distress"[tiab] OR "emotional disorder"[tiab] OR suicid*[tiab] OR "Mental Disorders"[Mesh] OR "Mental Health"[Mesh] OR "Stress, Psychological"[Mesh] OR "Psychological Distress"[Mesh] OR "Suicide"[Mesh]) AND (insecur*[tiab] OR secur*[tiab] OR precarity[tiab] OR "housing strain*"[tiab] OR "housing stress"[tiab] OR "housing crisis"[tiab] OR affordab*[tiab] OR unaffordab*[tiab] OR cost*[tiab] OR arrear*[tiab] OR "payment problem*"[tiab] OR "financial strain"[tiab] OR mobility[tiab] OR instab*[tiab] OR stability[tiab] OR stable[tiab] OR unstable[tiab] OR eviction[tiab] OR "multiple moves"[tiab] OR "frequent moves"[tiab] OR "forced moves"[tiab] OR "home loss"[tiab] OR "housing loss"[tiab] OR "duration of stay"[tiab] OR "doubling up"[tiab] OR overcrowding[tiab] OR "Housing Instability"[Mesh]) | ((TI (rent OR renter# OR rented OR renting OR rental OR tenan* OR "public housing" OR “social housing” OR “housing assistance” OR “housing subsid*” OR “housing allowance#” OR “housing benefit#” OR “subsidi?ed housing”)) OR (AB (rent OR renter# OR rented OR renting OR rental OR tenan* OR "public housing" OR “social housing” OR “housing assistance” OR “housing subsid*” OR “housing allowance#” OR “housing benefit#” OR “subsidi?ed housing”)) OR (KW (rent* OR tenan* OR "public housing" OR “social housing” OR “housing assistance” OR “housing subsid*” OR “housing allowance#” OR “housing benefit#” OR “subsidi?ed housing”))) AND ((TI (mental OR psych* OR "well#being" OR depress* OR “mood disorder#” OR “affective disorder” OR “bipolar disorder#” OR “emotional disorder” OR Anxiety OR “neurotic disorder#” OR schizo* OR “delusional disorder#” OR “somatoform disorder#” OR “stress#related disorder#” OR “stress disorder#” OR “Obsessive#compulsive” OR “trauma disorder#” OR “dissociative disorder#” OR “somatic symptom disorder#” OR “eating disorder#” OR “sleep#wake disorder#” OR “sexual dysfunction#” OR “paraphilic disorder#” OR “personality disorder#” OR “gender dysphoria” OR “disruptive disorder#” OR “impulse#control disorder#” OR “conduct disorder#” OR “behavio#r* disorder#” OR “substance#related disorder#” OR “addictive disorder#” OR “substance use” OR “neurocognitive disorder#” OR suicid*)) OR (AB (mental OR psych* OR "well#being" OR depress* OR “mood disorder#” OR “affective disorder” OR “bipolar disorder#” OR “emotional disorder” OR Anxiety OR “neurotic disorder#” OR schizophrenia OR schizotypal OR “delusional disorder#” OR “somatoform disorder#” OR “stress#related disorder#” OR “stress disorder#” OR “Obsessive#compulsive” OR “trauma disorder#” OR “dissociative disorder#” OR “somatic symptom disorder#” OR “eating disorder#” OR “sleep#wake disorder#” OR “sexual dysfunction#” OR “paraphilic disorder#” OR “personality disorder#” OR “gender dysphoria” OR “disruptive disorder#” OR “impulse#control disorder#” OR “conduct disorder#” OR “behavio#r* disorder#” OR “substance#related disorder#” OR “addictive disorder#” OR “substance use” OR “neurocognitive disorder#” OR suicid*)) OR (DE ("Well Being" OR "Subjective Well Being" OR "Mental Health" OR "Emotional Health" OR "Psychological Stress" OR "Psychiatric Symptoms" OR "Psychological Consequence" OR "Psychological Stress" OR "Psychopathology" OR "Psychosocial Factors" OR "Psychosocial Outcomes" OR "Mental Disorders" OR "Affective Disorders" OR "Disruptive Mood Dysregulation Disorder" OR "Major Depression" OR "Seasonal Affective Disorder" OR "Anxiety Disorders" OR "Castration Anxiety" OR "Generalized Anxiety Disorder" OR "Obsessive Compulsive Disorder" OR "Panic Attack" OR "Panic Disorder" OR "Phobias" OR "Separation Anxiety Disorder" OR "Trichotillomania" OR "Bipolar Disorder" OR "Borderline States" OR "Chronic Mental Illness" OR "Dissociative Disorders" OR "Eating Disorders" OR "Gender Dysphoria" OR "Mental Disorders due to General Medical Conditions" OR "Neurocognitive Disorders" OR "Neurodevelopmental Disorders" OR "Neurosis" OR "Paraphilias" OR "Personality Disorders" OR "Psychosis" OR "Serious Mental Illness" OR "Sleep Wake Disorders" OR "Somatoform Disorders" OR "Stress and Trauma Related Disorders" OR "Substance Related and Addictive Disorders" OR "Thought Disorders" OR "Anxiety" OR "Depression (Emotion)" OR "Suicidal Behavior" OR "Attempted Suicide" OR "Suicidal Ideation" OR "Suicide"))) AND ((TI (insecur* OR secur* OR precarity OR “housing strain#” OR “housing stress” OR “housing crisis” OR affordab* OR unaffordab* OR cost# OR arrear# OR “payment problem#” OR “financial strain” OR mobility OR instab* OR stability OR stable OR unstable OR eviction OR "multiple moves" OR "frequent moves" OR "forced moves" OR “home loss” OR "housing loss" OR “duration of stay” OR “doubling up” OR overcrowding)) OR (AB(insecur* OR secur* OR precarity OR “housing strain#” OR “housing stress” OR “housing crisis” OR affordab* OR unaffordab* OR cost# OR arrear# OR “payment problem#” OR “financial strain” OR mobility OR instab* OR stability OR stable OR unstable OR eviction OR "multiple moves" OR "frequent moves" OR "forced moves" OR “home loss” OR "housing loss" OR “duration of stay” OR “doubling up” OR overcrowding)) OR (KW (insecur* OR secur* OR precarity OR “housing strain#” OR “housing stress” OR “housing crisis” OR affordab* OR unaffordab* OR cost# OR arrear# OR “payment problem#” OR mobility OR instab* OR stability OR stable OR unstable OR eviction OR "multiple moves" OR "frequent moves" OR "forced moves" OR “home loss” OR "housing loss" OR “duration of stay” OR “doubling up” OR overcrowding))) | TS = (rent* OR tenan* OR "public housing" OR “social housing” OR “housing assistance” OR “housing subsid*” OR “housing allowance$” OR “housing benefit$” OR “subsidi$ed housing”) AND TS = (mental OR psych* OR "well$being" OR depress* OR “mood disorder$” OR “affective disorder” OR “bipolar disorder$” OR “emotional disorder$” OR Anxiety OR “neurotic disorder$” OR schizo* OR “delusional disorder$” OR “somatoform disorder$” OR “stress*related disorder$” OR “stress disorder$” OR “obsessive$compulsive” OR “trauma disorder$” OR “dissociative disorder$” OR “somatic symptom disorder$” OR “eating disorder$” OR “sleep*wake disorder$” OR “sexual dysfunction$” OR “paraphilic disorder$” OR “personality disorder$” OR “gender dysphoria” OR “disruptive disorder$” OR “impulse*control disorder$” OR “conduct disorder$” OR “behavi$r* disorder” OR “substance*related disorder$” OR “addictive disorder$” OR “substance use” OR “neurocognitive disorder$” OR suicid*) AND TS = (insecur* OR secur* OR precarity OR “housing strain$” OR “housing stress” OR “housing crisis” OR affordab* OR unaffordab* OR cost$ OR arrear$ OR “payment problem$” OR “financial strain” OR mobility OR instab* OR stability OR stable OR unstable OR eviction OR "multiple moves" OR "frequent moves" OR "forced moves" OR “home loss” OR "housing loss" OR “duration of stay” OR “doubling up” OR overcrowding) | noft(rent* OR tenan* OR "public housing" OR “social housing” OR "housing assistance" OR "housing subsid*" OR "housing allowance*" OR "housing benefit*" OR "subsidi?ed housing" OR "Housing assistance programmes") AND noft(mental OR psych* OR "well*being" OR depress* OR "mood disorder*" OR "affective disorder" OR "bipolar disorder*" OR “emotional disorder” OR Anxiety OR "neurotic disorder*" OR schizo* OR "delusional disorder*" OR "somatoform disorder*" OR "stress*related disorder*" OR "stress disorder*" OR "Obsessive*compulsive" OR "trauma disorder*" OR "dissociative disorder*" OR "somatic symptom disorder*" OR "eating disorder*" OR "sleep*wake disorder*" OR "sexual dysfunction*" OR "paraphilic disorder*" OR "personality disorder*" OR "gender dysphoria" OR "disruptive disorder*" OR "impulse*control disorder*" OR "conduct disorder*" OR “behavio*r disorder*” OR "substance*related disorder*" OR "addictive disorder*" OR "substance use" OR "neurocognitive disorder*" OR “emotional distress” OR suicid*) AND noft(insecur* OR secur* OR precarity OR "housing strain*" OR "housing stress" OR "housing crisis" OR affordab* OR unaffordab* OR cost* OR arrear* OR "payment problem*" OR mobility OR Instab* OR stability OR stable OR unstable OR eviction OR "multiple moves" OR "frequent moves" OR "forced moves" OR "home loss" OR "housing loss" OR "duration of stay" OR "doubling up" OR overcrowding) |
| **Results, 09^th^ December 2022** | | | **444** | **365** | **775** | **203** |
