## Supplementary material for "Exploring the association between housing insecurity and mental health among renters: A systematic review": S5_Appendix

**S5 Appendix. Reference List of the Measurement Tools Used in the Included Studies.**

Barry, K. L., & Fleming, M. F. (1993). The alcohol use disorders identification test (AUDIT) and the SMAST-13: predictive validity in a rural primary care sample. *Alcohol and Alcoholism*, *28*(1), 33–42. https://doi.org/10.1093/oxfordjournals.alcalc.a045346

Cook, W. W., & Medley, D. M. (1954). Proposed hostility and Pharisaic-virtue scales for the MMPI. *Journal of Applied Psychology*, *38*(6), 414–418. https://doi.org/10.1037/h0060667

Derogatis, L. R. (1993). *Brief symptom inventory: BSI*. Pearson. https://doi.org/10.1037/t00789-000

Endicott, J., Nee, J., Harrison, W., & Blumenthal, R. (1993). Quality of Life Enjoyment and Satisfaction Questionnaire: A new measure. *Psychopharmacology Bulletin*, *29*(2), 321–326.

Goldberg, D. P., & Williams, P. (1988). *A user’s guide to the General Health Questionnaire*. NFER-Nelson.

Kessler, R. C., Andrews, G., Colpe, L. J., Hirip, E., Mroczek, D. K., Normand, S. L. T., Walters, E. E., & Zaslavsky, A. M. (2002). Short screening scales to monitor population prevalences and trends in non-specific psychological distress. *Psychological Medicine*, *32*(6), 959–976. https://doi.org/10.1017/s0033291702006074

Kessler, R. C., Andrews, G., Mroczek, D., Ustun, B., & Wittchen, H.‑U. (1998). The World Health Organization Composite International Diagnostic Interview short-form (CIDI-SF). *International Journal of Methods in Psychiatric Research*, *7*(4), 171–185. https://doi.org/10.1002/mpr.47

Kroenke, K., & Spitzer, R. L. (2002). The PHQ-9: A New Depression Diagnostic and Severity Measure. *Psychiatric Annals*, *32*(9), 509–515. https://doi.org/10.3928/0048-5713-20020901-06

Kroenke, K., Spitzer, R. L., & Williams, J. B. W. (2003). The Patient Health Questionnaire-2: Validity of a two-item depression screener. *Medical Care*, *41*(11), 1284–1292. https://doi.org/10.1097/01.MLR.0000093487.78664.3C

Kroenke, K., Spitzer, R. L., Williams, J. B. W., & Löwe, B. (2009). An ultra-brief screening scale for anxiety and depression: The PHQ-4. *Psychosomatics*, *50*(6), 613–621. https://doi.org/10.1176/appi.psy.50.6.613

Kroenke, K., Spitzer, R. L., Williams, J. B. W., Monahan, P. O., & Löwe, B. (2007). Anxiety disorders in primary care: Prevalence, impairment, comorbidity, and detection. *Annals of Internal Medicine*, *146*(5), 317–325. https://doi.org/10.7326/0003-4819-146-5-200703060-00004

Löwe, B., Gräfe, K., Zipfel, S., Spitzer, R. L., Herrmann-Lingen, C., Witte, S., & Herzog, W. (2003). Detecting panic disorder in medical and psychosomatic outpatients: Comparative validation of the Hospital Anxiety and Depression Scale, the Patient Health Questionnaire, a screening question, and physicians' diagnosis. *Journal of Psychosomatic Research*, *55*(6), 515–519. https://doi.org/10.1016/s0022-3999(03)00072-2

Radloff, L. S. (1977). The CES-D Scale: A Self-Report Depression Scale for Research in the General Population. *Applied Psychological Measurement*, *1*(3), 385–401. https://doi.org/10.1177/014662167700100306

Tennant, R., Hiller, L., Fishwick, R., Platt, S., Joseph, S., Weich, S., Parkinson, J., Secker, J., & Stewart-Brown, S. (2007). The Warwick-Edinburgh Mental Well-being Scale (WEMWBS): Development and UK validation. *Health and Quality of Life Outcomes*, *5*(1), 63. https://doi.org/10.1186/1477-7525-5-63

Ware, J. E. (1993). *SF-36 health survey: Manual and interpretation guide*. The Health Institute, New England Medical Center.

Ware, J. E., Kosinski, M., Tuner-Bowker, D. M., & Gandek, B. (2002). *Version 2 of the SF-12 health survey*. Quality Metric Inc.

Weathers, F. W., Litz, B. T., Keane, T. M., Palmieri, P. A., Marx, B. P., & Schnurr, P. P. (2003). *The PTSD Checklist for DSM-5 (PCL-5)*. Scale available from the National Center for PTSD at www.ptsd.va.gov.
